# Correlations of Mobility and Covid-19 Transmission in Global Data

**DOI:** 10.1101/2020.05.06.20093039

**Authors:** Nittai K. Bergman, Ram Fishman

## Abstract

Assessing the contribution of mobility declines to the control of Covid-19 diffusion is an urgent challenge of global import. We analyze the temporal correlation between transmission rates and societal mobility levels using daily mobility data from Google and Apple in an international panel of 99 countries and a panel of all states in the United States. Reduced form regression estimates that flexibly control for time trends suggest that a 10 percentage point reduction in mobility is associated with a 0.04-0.07 reduction in the value of the effective reproduction number, *R*(*t*), depending on geographical region and modelling choice. According to these estimates, to avoid the critical value of *R* = 1, easing mobility restrictions may have to be limited or delayed until other non-mobility related preventative measures reduce *R* to a level of approximately 0.7 in Europe, 0.75 in Asia, and 0.8 in the United States. Given gaps in data availability and inference challenges, these estimates should be interpreted with caution.

## Introduction

By some estimates, more than a third of the global population have been subjected to severe mobility restrictions since the start of the Covid-19 pandemic. Numerous governments around the world have resorted to such “lockdowns” as their primary strategy of limiting the transmission of infection, at enormous economic and social costs. As costs escalate, and transmission rates decline in some countries, a growing debate has emerged regarding when and how lockdowns should be eased, and whether it is possible to do so without unleashing additional waves of infection. An assessment of the relation between mobility levels and transmission rates can be of value in helping to navigate this policy dilemma and in understanding the determinants of diffusion.

Several papers have estimated the declines in transmission rates that occurred following lockdowns and other non pharmaceutical interventions (NPI) by using detailed case-level data in specific localities or countries—mainly Wuhan and France (see., e.g., Leung et al., 2020; Lipsitch et al., 2020; Pan et al., 2020; Wang et al., 2020; Salje et al., 2020; Roux et al., 2020). Hsiang et al. (2020) use detailed sub-national panel data to estimate the impacts of NPIs in China, South Korea, Iran, Italy, France, and the United States (US) and find variable, but overall large impacts on transmission rates. Flaxman (2020a; 2020b) and Kučinskas (2020) estimate the effects of NPIs on effective reproduction numbers using a panel of 14 European states, finding that the results are sensitive to the manner in which the regression model is specified.

In this paper, we use publicly available data to empirically estimate the relation between transmission rates (effective reproduction numbers) and societal mobility levels using a large, international 99-country panel, as well as a panel covering all states in the United States, covering the period between late February and early May 2020. Our assessment employs a reduced-form regression analysis based on daily mobility data provided by Google and Apple, estimates of daily transmission rates at the country level from Kučinskas (2020), and daily estimates of transmission rates at the U.S. state level from Systrom and Vladeck (2020).^4^

While studying the effect of lockdowns is clearly important, we focus on the association between transmission rates and mobility rather than on the association between transmission rates and government NPIs, and in particular, lockdown orders. \\We do so for three reasons. The first is that evidence suggests an imperfect correspondence between lockdowns and mobility levels, with mobility declining prior to lockdowns (or even in their absence), and in certain cases increasing prior to formal lockdown easing (see e.g., Gupta et al., 2020). Second, as governments consider the degree to which lockdown conditions can be eased, it is important to analyze the span of the relation between mobility and transmission rates and not only the effects of discrete, large reductions in mobility resulting from lockdown orders.^5^ Finally, lockdowns may impact transmission rates not solely due to their direct effect on mobility rates, but also through their impact on other forms of individual behavior. Future changes in mobility levels need not necessarily come about in tandem with such changes in individual level behavior. One additional analysis of the relation between mobility and Covid-19 transmission rates is provided by Kissler et al. (2020) who show that reductions in between-borough commuting movements in New York City are negatively correlated with Covid-19 prevalence.

A visual inspection of the country level data suggests mixed patterns regarding the relation between transmission rates and mobility. In Australia, for example (Figure 1, top panel), transmission rates seem to track mobility levels: during the initial sample period, steep reductions in mobility are followed by significant reductions in transmission rates, while subsequent to a rise in mobility commencing mid-April, transmission rates rise as well. In contrast, in Germany (Figure 1, middle panel) transmission rates fall following steep reductions in mobility levels, but they continue their decline even after mobility levels start to rise at the end of March. In South Korea (Figure 1, bottom panel), transmission rates decline significantly by the beginning of March, and only initially coincide with relatively modest reductions in mobility, implying a weaker relation between the two.

**Figure 1:**
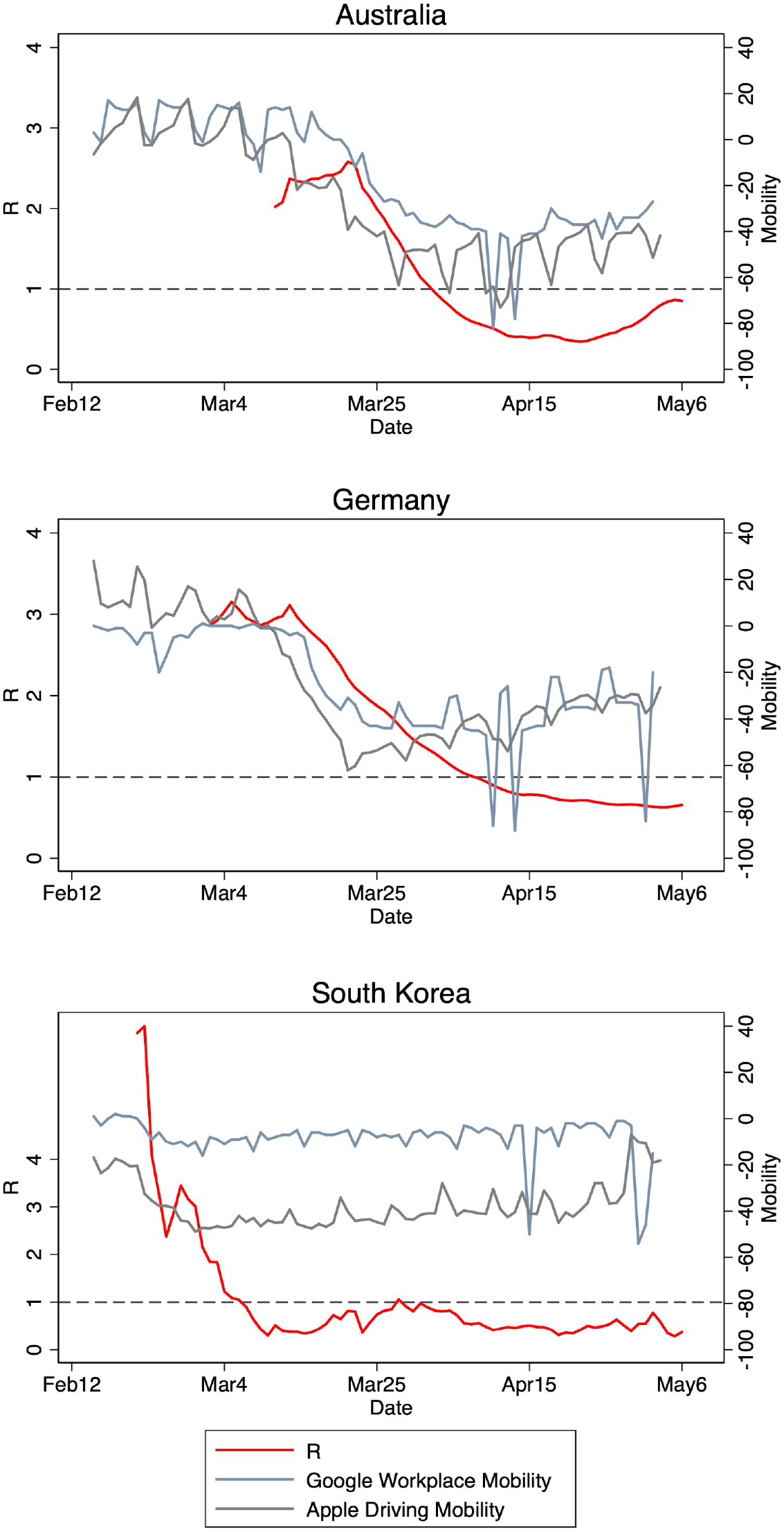
Plots of estimated effective reproduction numbers *R*(*t*), Google workplace mobility and Apple driving mobility indicators for Spain and South Korea. *R*(*t*) is plotted against the left axis. Mobility changes from baseline (in percentage points, see data section) are plotted against the right axis.

Our reduced form regression analysis is designed to estimate the average correlation between mobility and transmission rates in a large international sample of countries. By providing such summary estimates at global scale, our analysis complements important studies that analyze sub-national transmission dynamics and underlying epidemiological processes at finer resolution.

As discussed in detail below, there are significant challenges in estimating and interpreting the transmission-mobility relation. First, the limited precision of transmission indicators may introduce substantial measurement error into the data, making it hard to identify a correlation. Second, changes in mobility may also endogenously respond to infection rates. Third, variation in mobility levels may be correlated with variation in other forms of preventative behavior, whether voluntary or government dictated, meaning that any observed correlation can be wrongfully attributed to changes in mobility levels.

Given the absence of detailed data on such behavior, we employ standard panel data techniques to absorb some of the variation in these unobserved potential confounders. In particular, we base our estimates only on variation in mobility and transmission that occur within countries over time, rather than on variation across countries, which is especially prone to omitted variable bias (e.g. economic development levels, infrastructure, health system functionality). Further, we include various forms of flexible temporal trends in our regressions to capture some of the temporal variation in potential confounders that may occur within countries over time. Nevertheless, we emphasize that a great deal of caution must be exercised in interpreting our estimates as capturing the causal impacts of mobility changes on transmission. For that reason, we also emphasize that there is no straightforward way to infer policy prescriptions from the correlations we estimate, as policy induced changes in mobility may impact other variables which influence transmission rates. As such, our results do not provide definitive conclusions, but should be viewed as a first step that utilizes publicly available measures of mobility levels and transmission to study a question of enormous social import.

Our current point estimates from the international cross-country panel suggest that a 10 percentage point (p.p.) reduction in mobility is associated with a 0.05-0.07 reduction in the value of the effective reproduction number, *R*(*t*), depending on geographical region and modelling choice. Estimates from the U.S. state-level panel suggest that a 10 p.p. reduction in mobility is associated with roughly a 0.04 reduction in the value of the effective reproduction number, *R*(*t*).

Our analysis documents a decline in the strength of the mobility-transmission correlation over the study period. During the month of March, mobility levels exhibit dramatic declines in many countries in the world. During this intense “lockdown” period, we estimate that a 10 p.p. reduction in mobility is associated with a 0.06-0.09 reduction in *R*(*t*). In April, in contrast, the estimated association remains positive, but drops by roughly a factor of three to 0.02-0.03 units of *R*(*t*). This decline could potentially reflect fundamental differences between the dynamics of the initial “lockdown” and the later phase of the crisis driven by individual-level changes in behavior (such as individual level social distancing, hygiene, mask use) or government-level steps to curb transmission. In addition, changes in environmental conditions may also affect transmission rates (see, e.g., Carleton et al., 2020) in a manner which weakens the association between mobility and Covid-19 transmission.

With the appropriate caveats in mind, our estimates of the relation between mobility and transmission rates imply that in order to avoid allowing *R*(*t*) to exceed unsustainable levels, easing of mobility restrictions may have to be limited or delayed until further declines in *R* are achieved through other means.^6^ Indeed, assuming an additive model for the impact of mobility and non-mobility related suppression methods on transmission rates, to avoid the critical value of *R =* 1, our estimates suggest that *R*(*t*) may have to be reduced to levels of about 0.55–0.7 in Europe, 0.64–0.76 in Asia, and approximately 0.8 in the United States before mobility levels are fully restored to pre-pandemic levels.^7^

## Data and Methods

### Mobility Data

The principal measure of mobility used in this analysis is taken from the *Covid 19 Community Mobility Reports* provided by Google. As an alternative measure, we also use the *Mobility Trends Reports* provided by Apple. The Google data utilize anonymized location-based information to assess changes in the number of visits to several categories of locations in a given day and country, as compared to a baseline value for that day of week.^8^ The categories include retail and recreation, groceries and pharmacy, parks, transit stations, workplaces, and residential.^9^ Similarly, the Apple data report the “relative volume of directions requests” sent using the Apple Maps application, compared to a baseline volume on January 13th, 2020. The Apple data differentiate between three types of direction requests: driving, walking, and transit. Google data is available for 132 countries, whereas Apple data is available for 63 countries. Figure S1 plots Google data on visits to workplaces over time, averaged in six world regions. Figure S2 plots the six measures of Google mobility data over time, averaged over Europe.

Both Google and Apple data are updated continuously. Google data are available for the period February 15th–May 2nd and Apple data for the period January 13th-May 3rd.^10^

### Covid-19 Transmission Rates

The preferred indicator of Covid-19 transmission rates is the effective reproduction number *R*(*t*), which measures the number of individuals an average infected person infects during the period of infection. In the primary analysis, we make use of two independent sets of estimates of *R*(*t*), one at the country level and one for U.S. states.

Country level estimates of *R*(*t*) are provided by Kučinskas (2020) between January 23rd and May 6th for 124 countries (temporal coverage varies by country and begins after 100 cases are confirmed). To construct this proxy, Kučinskas (2020) uses data on new cases, recoveries, and deaths and backs out estimates of *R*(*t*) on the basis of disease models. Importantly, the data (and the estimates) are smoothed with Kalman-filtering techniques. This means that discrete, high frequency movements in the actual effective reproduction number will be difficult to observe in these estimates. In addition, the estimates do not account for the delay between actual infection and official diagnosis. As such, they reflect lagged infection rates, with a lag size that combines the delay between infection and Covid-19 testing and the time between testing and official reporting of test results. In our analysis we assume an overall lag of seven days to account for an incubation period of approximately 4-5 days (Qun Li et al. 2020) and a 2-3 day lag between testing and the official recording of positive test results.^11^

As is well known, a major limitation shared by all proxies based on confirmed case counts is that they are likely to substantially underestimate the true number of cases in the population. To the extent that the ratio of confirmed to actual cases is constant within countries (even if not between countries), however, this will not bias the *R* estimates. Further, Kučinskas (2020) argues that the estimation method is “robust in the sense that the estimates of *R* remain fairly accurate even when new cases are imperfectly measured, or the true dynamics of the disease do not follow the SIR model”.

For the state-level analysis in the United States, estimates of *R*(*t*) at the state level are provided by Sysrom and Vladeck (2020), which adjusts the estimates for state-level testing capacity and for the delay in test reporting.

### Sample

The sample studied in our international analysis includes 4,804 observations from 99 countries (shown in Figure S3) over the period March 13th - May 2nd when using Google data, and 3,294 observations from 61 countries over the period March13th - May 8th when using Apple data. In both cases, data coverage is uneven across countries, starting when the confirmed number of cases reaches 100 in each country.

Figure S4 plots our principal mobility measure (Google visits to workplaces) and estimates of *R*(*t*) against time for those countries that have more than 30 days in which both data exist.

The U.S. sample includes all states over the period March 13th - May 2nd when using Google data, and March 13th - May8th when using Apple data.

### Empirical Strategy

We employ standard panel-regression techniques to estimate the association between transmission rates (proxied by the *R* estimates described above) and mobility measures. The regressions include country specific fixed effects (intercepts) to flexibly account for all time-invariant country attributes, thus basing estimates of the relation between transmission rates and mobility on the correlation between these two variables over time within countries.^12^

In addition to the inherent limitations of the precision with which transmission rates are measured, a significant challenge in estimating the transmission-mobility relationship is the likely presence of time varying behavioral, environmental, and epidemiological variables that can also affect transmission rates and which are difficult to observe. Behavioral variables can include hygienic practices, mask usage rates, and dimensions of social distancing not captured by mobility (such as, for example, maintaining minimal distance between individuals and maximal room occupancy rates). Epidemiological models predict transmission rates to respond to the diffusion of the virus in the population. As one example, in standard SIR models, effective reproduction numbers rates decline over time with the fraction of susceptible individuals in the population. Environmental factors may include temperature, UV radiation, and humidity (Carleton et al, 2020). Failing to control for these variables in the regressions may reduce the precision of the estimates, and may also bias them whenever these variables are correlated with mobility and affect transmission rates.

An additional concern in analyzing the relation between infection rates and mobility levels is the endogenous nature of mobility behavior. In particular, mobility rates are influenced by individual choice as well as by policy directives (such as lockdown easing and tightening), which may in turn be influenced by disease transmission rates and rising case counts.

For the above reasons, panel-regression estimates relating within-country variation in infection rates to levels of mobility may be subject to bias and should be interpreted with a good deal of caution. Unless all potential confounders are observed, or exogenous variation in mobility is utilized, the estimates are not amenable to causal interpretation. We note, however, with the appropriate caution, that many of the unobserved confounders would tend to bias our estimates on the relation between mobility and transmission rates upwards, as variation in these confounders likely served to reduce transmission rates in tandem with mobility restrictions (examples include increased mask usage and hygiene).

To partially alleviate concerns stemming from unobservable within-country time variation, our analysis includes a host of fixed effects and time trends that may capture some of the unobserved variation. As mentioned above, all regressions estimated include country (or state) specific fixed effects. The regressions also flexibly control for temporal trends using individual date fixed effects, at either the global or regional level.^13^ Such fixed effects absorb the potentially confounding influences of any time varying factor that behaves in a similar manner globally or regionally. They also help absorb variation stemming from within-week mobility cycles. In another check, we flexibly control not for calendar time, but for fixed effects of the number of days which have elapsed in each country since the 100th case was confirmed. This allows us to absorb the potentially confounding effects of, for example, dynamical factors related to the ecological evolution of the pandemic in the absence of interventions. In a final test, we allow the regressions to include a separate (linear) time-trend for each country. We remain acutely aware, however, that none of these approaches can fully address the possibility that our results are biased by unobserved confounders, and emphasize the need for caution in interpreting them.

Formally, we estimate the following baseline regression:

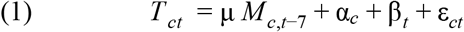

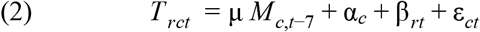

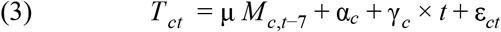

where *T* is a proxy for Covid-19 transmission rates in country *c* (in region *r)* on date *t* as described above, and *M* is one of the mobility measures described above, measured seven days before the date at which *T* is observed. The regression include country fixed effects α*_c_*, date fixed effects β*_t_* at either global (eq.1) or regional (eq.2) levels, or country specific time trends (eq.3). Because transmission and mobility may exhibit temporal autocorrelation within countries, all standard errors are clustered at the country level. For the U.S. panel, we run similar regressions to (1) and (3) except that the unit of observation is the state-day.

As explained above, when analyzing the international panel, we use 7-day lagged values of mobility in the regression to account for the delay between infection and confirmation of the case. Estimates of *R*(*t*) used in the U.S. analysis already account for this delay, so we use the contemporaneous value. Tests of alternative lag periods, displayed in Figure S5, provide support for this choice: when lagged values of between 0-3 weeks are included in the regression, the lags we choose in practice show the largest and more significant coefficients.

Finally, since mobility measures are strongly correlated temporally within countries (Figure S2), separating out the effects of each type of mobility indicator demands statistical power that is unlikely to be provided by the current sample. Our main regression models therefore include a single measure of societal mobility as the explanatory variable. We focus on the Google workplace mobility measure, being a natural proxy for economic activity. In an exploratory analysis reported below, we also estimate models that simultaneously include the various components of mobility levels (workplace, residential, transit, etc).

## General Trends in Mobility and Transmission

During the earlier part of the sample period, the data exhibit significant downward trends in both mobility and estimated transmission rates. Figure 2 (top panel) exhibits the daily average of Google workplace mobility, Apple driving-based mobility, and (unlagged) *R*(*t*) estimates over the sample period in Europe. As can be seen, average European mobility levels decline sharply between mid-February and mid-March (with Google workplace mobility declining by approximately 50 percentage points), stabilize between mid-March and mid-April, and subsequently display a rising trend. Over the same time period, Figure 2 (top panel) shows a decline in estimated *R* values from 3.5 to below approximately 0.8.

**Figure 2:**
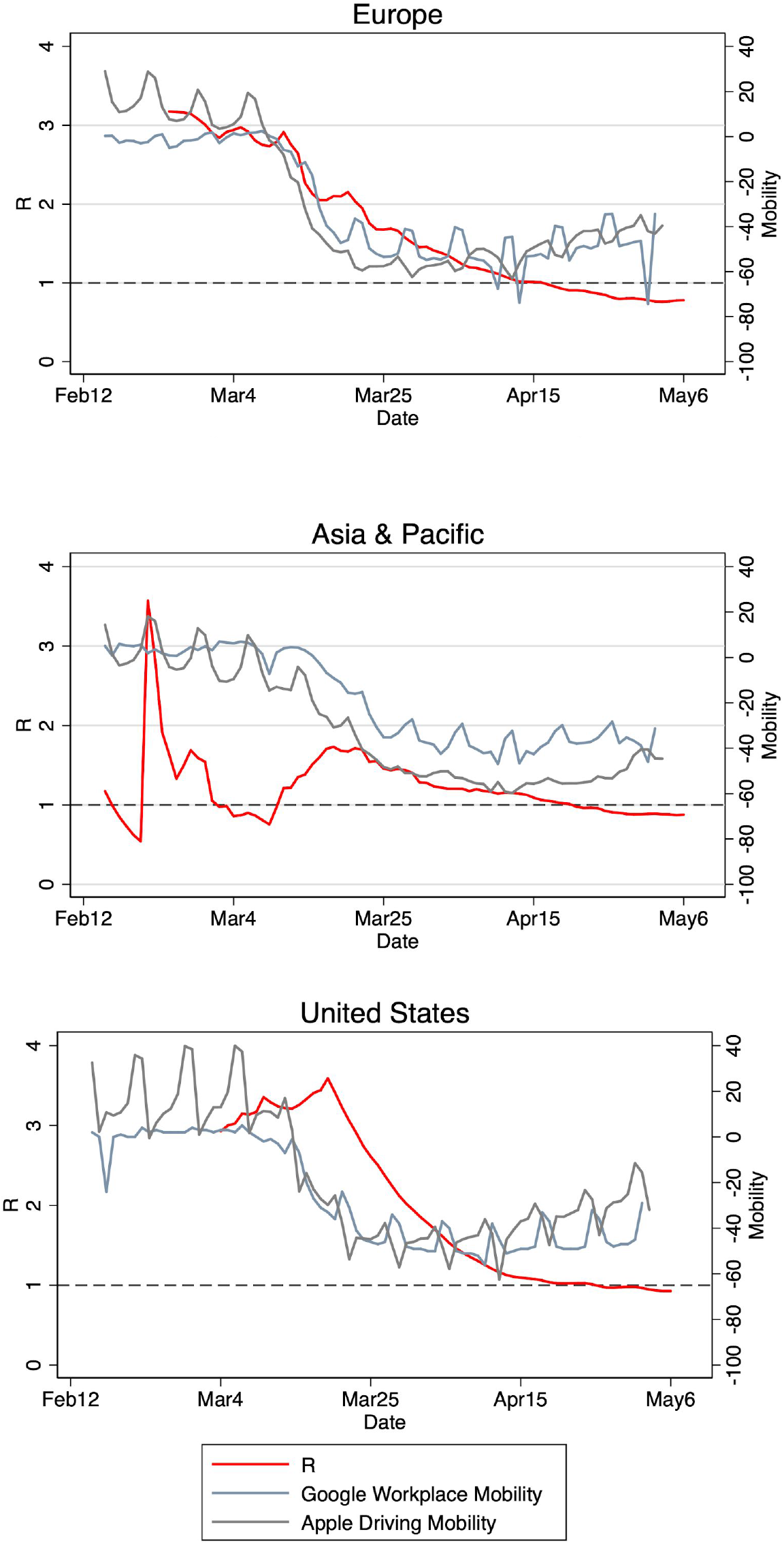
Plots of *R*(*t*), Google (visit to workplaces) and Apple (driving searches) mobility indicators over time, averaged for Europe and Asia. *R*(*t*) is plotted against the left axis. Mobility changes from baseline (in percentage points, see data section) are plotted against the right

Figure 2 (middle panel) provides analogous information for countries in the sample within the “Asia and Pacific” region. Similar to Europe, the figure indicates a decline of average *R* of approximately 3.1 units from a peak of 4 to a value of 0.9, and a decline of 40 percentage points in the Google workplace mobility measure. The bottom panel of Figure 2 focuses on the United States using the state-level panel dataset. As can be seen, the average reproduction number falls by approximately 0.50 units during the sample period (from approximately 1.4 to 0.9), while the average Google workplace mobility levels decline by 45 percentage points. We note that the sample period of state-level reproduction numbers, *R*, begins only in mid-March, and so we cannot rule out higher levels of *R*—similar to those observed in Europe and Asia—prior to that.

Changes in average mobility levels mask a good deal of country-level heterogeneity. Figure S4 in the supplementary material plots Google workplace mobility measures and estimated *R* values for sixty countries that have at least 30 days of both mobility and transmission data.

Figure 3 (top panel) provides two snapshots of the international data, showing scatter plots of each country’s estimated reproduction number *R* against its level of Google workplace mobility on March 15th and May 1st. Between the two periods, there is clear movement of most countries towards lower reproduction numbers and mobility levels. There is no indication of a significant cross-sectional relation between *R* and mobility levels in either period.

**Figure 3:**
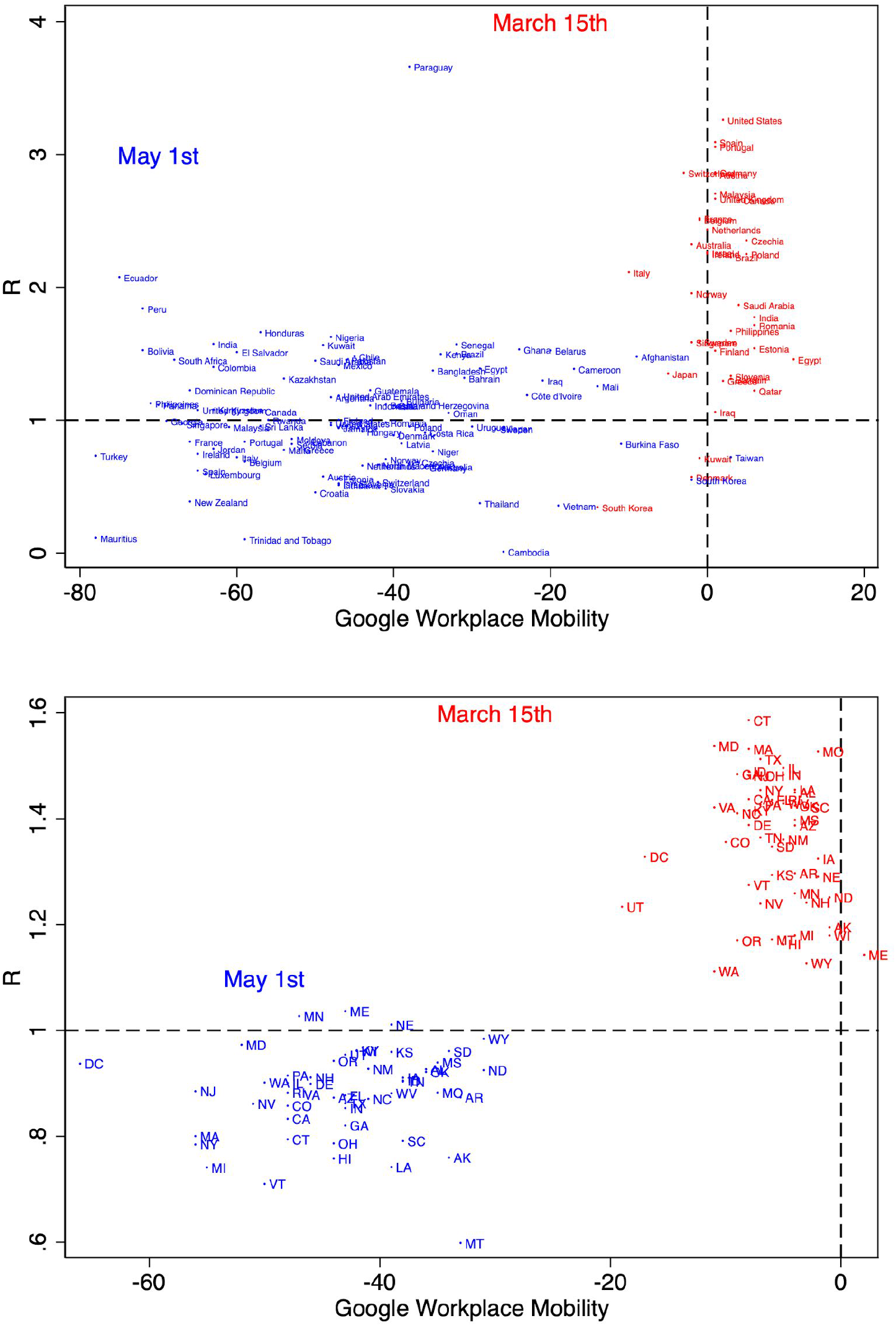
Scatter plots of estimated R values vs. Google workplace mobility for countries (top panel) and U.S. states (bottom panel) on March 15th (red) and May 1st (blue).

The bottom panel of Figure 3 provides a parallel graph for U.S. states on the same two dates. Here too, during the initial stage of the pandemic in the United States, transmission rates are high, mobility levels have not yet declined, and there is no discernable cross-sectional relation between the two. In contrast, by May 1st, transmission rates and mobility levels decline significantly across the United States, and display a positive correlation.

However, cross-sectional correlations of this kind can easily be confounded by numerous other sources of heterogeneity (even if such heterogeneity could be lower across U.S. states than across countries). It is for this reason that, in order to estimate the relation between R and mobility, the analysis below focuses on variation within countries over time.

## Results

We begin by reporting results from the international sample. The results are summarized in Figure 4, and tabulated in Tables 1-7.

**Figure 4:**
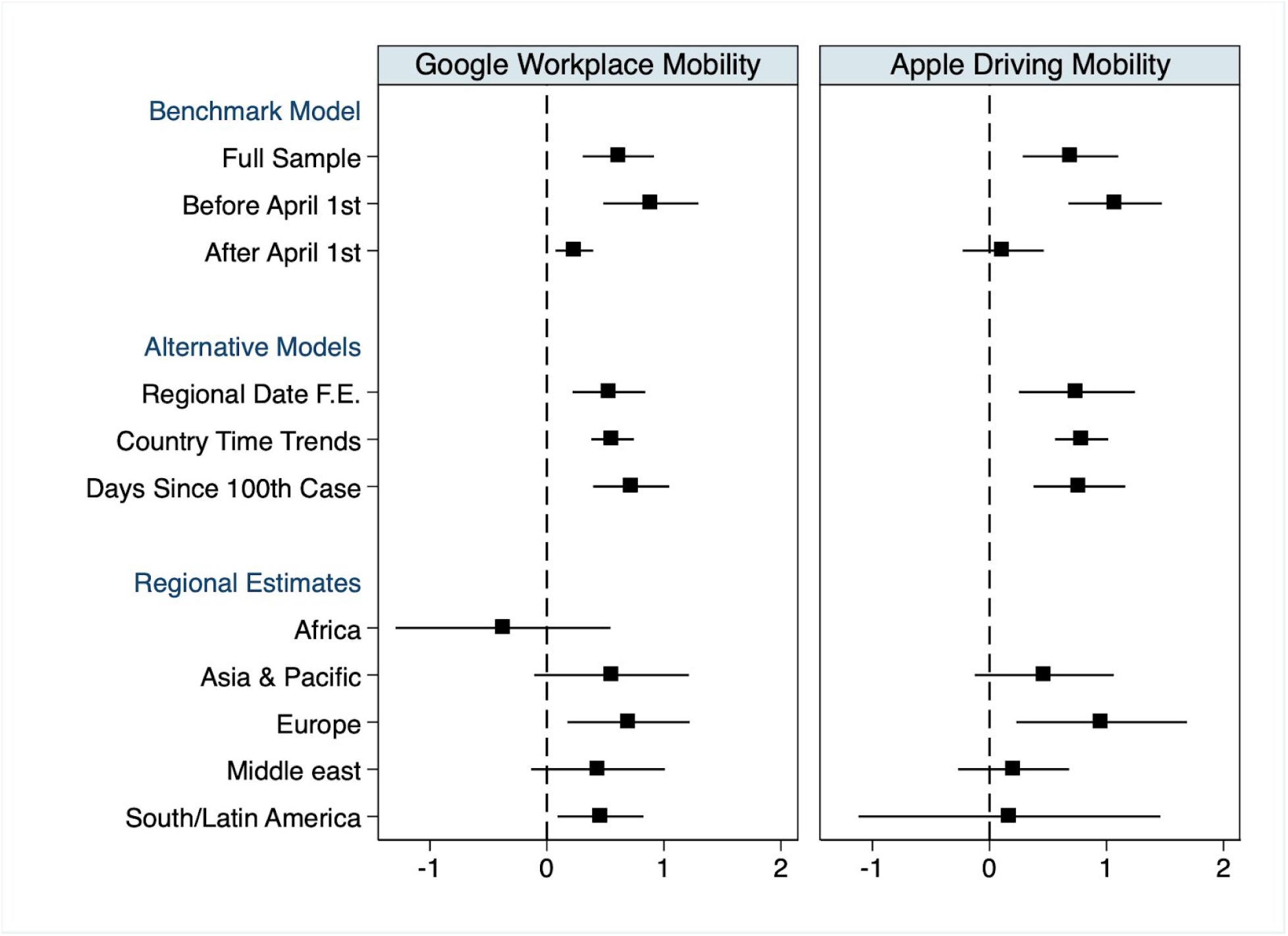
Coefficients from regressions of estimated effective reproduction numbers *R*(*t*) observed at the country-date level on Google workplace mobility (left panel) and Apple driving mobility (right panel) indicators (see text for details). Markers indicate point estimates and error bars indicate 95% confidence intervals corresponding to standard errors that are clustered at the country level. The benchmark model includes country and date fixed effects. Alternative models include additional controls as specified.

**Table 1:** Baseline regression results relating estimated *R* values to seven-day lagged Google workplace mobility. Each column reports results from a separate regression. All regressions include country fixed effects. Standard errors, clustered by country, are reported in parentheses. Stars indicate statistical significance (*p<0.1, **p<0.05, ***p<0.01).

|  | (1) | (2) | (3) | (4) |
| --- | --- | --- | --- | --- |
|  | **R** | **R** | **R** | **R** |
| Google Workplace Mobility<br>(Seven-day Lagged) | 0.61*** | 0.53*** | 0.56*** | 0.72*** |
|  | (0.15) | (0.16) | (0.09) | (0.16) |
| Observations | 4803 | 4779 | 4803 | 4802 |
| Adjusted R-squared | 0.640 | 0.729 | 0.813 | 0.650 |
| Date F.E. | Global | Regional | Global | N |
| Country Specific Time Trends | N | N | N | Y |
| Days Since 100th Case F.E. | N | N | Y | N |

**Table 2:** Regression results relating estimated *R* values to seven-day lagged Apple driving mobility. Each column reports results from a separate regression. All regressions include country fixed effects. Standard errors, clustered by country, are reported in parentheses. Stars indicate statistical significance (*p<0.1, **p<0.05, ***p<0.01).

|  | (1) | (2) | (3) | (4) |
| --- | --- | --- | --- | --- |
|  | **R** | **R** | **R** | **R** |
| Apple Driving Mobility<br>(Seven-day Lagged) | 0.69*** | 0.75*** | 0.79*** | 0.77*** |
|  | (0.20) | (0.25) | (0.11) | (0.20) |
| Observations | 3294 | 3228 | 3295 | 3293 |
| Adjusted R-squared | 0.727 | 0.777 | 0.839 | 0.727 |
| Date F.E. | Global | Regional | Global | N |
| Country Specific Time Trends | N | N | N | Y |
| Days Since 100th Case F.E. | N | N | Y | N |

**Table 3:** Regression results relating estimated *R* values (obtained from rt.live) to Google workplace mobility (Columns 1-2) and Apple driving mobility (Columns 3-4) measures in the U.S. panel. Each column reports results from a separate regression. All regressions include state fixed effects. Standard errors, clustered by country, are reported in parentheses. Stars indicate statistical significance (*p<0.1, **p<0.05, ***p<0.01).

|  | (1) | (2) | (3) | (4) |
| --- | --- | --- | --- | --- |
|  | **Google Workplace** |  | **Apple Driving** |  |
| Mobility | 0.36*** | 0.47*** | 0.13** | 0.32*** |
|  | (0.13) | (0.04) | (0.06) | (0.02) |
| Observations | 2601 | 2601 | 2850 | 2850 |
| Adjusted R-squared | 0.888 | 0.901 | 0.886 | 0.887 |
| Date F.E. | Y. | N | Y | N |
| StateSpecific<br>Time Trends | N | Y | N | Y |

**Table 4:** Regional regression results relating estimated *R* values to seven-day lagged Google workplace mobility. Each column report results from a separate regression. All regressions include country fixed effects. Standard errors, clustered by country, are reported in parentheses. Stars indicate statistical significance (*p<0.1, **p<0.05, ***p<0.01). Apple mobility data is not available in Africa.

|  | (1) | (2) | (3) | (4) | (5) |
| --- | --- | --- | --- | --- | --- |
|  | **R** | **R** | **R** | **R** | **R** |
|  | **Africa** | **Asia & Pacific** | **Europe** | **Middle East** | **South/ Latin America** |
|  | Google Workplace Mobility |  |  |  |  |
| Mobility (7-day Lagged) | -0.37 | 0.55* | 0.70** | 0.44 | 0.46** |
|  | (0.42) | (0.31) | (0.26) | (0.25) | (0.18) |
| Observations | 446 | 960 | 1966 | 512 | 783 |
| Adjusted R-squared | 0.436 | 0.606 | 0.794 | 0.487 | 0.814 |
|  | Apple Driving Mobility |  |  |  |  |
| Mobility (7-day Lagged) |  | 0.47 | 0.96** | 0.21 | 0.17 |
|  |  | (0.27) | (0.36) | (0.15) | (0.50) |
| Observations |  | 719 | 1905 | 200 | 292 |
| Adjusted R-squared |  | 0.605 | 0.820 | 0.565 | 0.910 |
| Date Fixed Effects | Regional | Regional | Regional | Regional | Regional |

**Table 5:** Regression results relating estimated *R* values to all seven-day lagged Google mobility indicators, jointly estimated. Dependent variable: estimates of *R* from Kučinskas (2020) in Columns 1,2,4,5 and from rt.live in Column 3. Each column reports results from a separate regression. Columns 1,2,3: Google mobility indicators. Column 4,5: Apple mobility indicators, with all mobility indicators lagged seven days (other than in Column 3). All regressions include country fixed effects (column 3 includes state fixed effects). Standard errors, clustered by country (or state), are reported in parentheses. Stars indicate statistical significance (*p<0.1, **p<0.05, ***p<0.01).

|  | (1) | (2) | (3) |  | (4) | (5) |
| --- | --- | --- | --- | --- | --- | --- |
|  | **R** | **R** | **R** |  | **R** | **R** |
| **Google Mobility** |  |  |  | **Apple Mobility** |  |  |
| **Retail and Recreation** | 1.03*** | 0.66** | 0.17 |  |  |  |
|  | (0.34) | (0.29) | (0.16) |  |  |  |
| **Grocery and Pharmacy** | -0.40*** | -0.28* | -0.08 |  |  |  |
|  | (0.14) | (0.15) | (0.10) |  |  |  |
| **Parks** | -0.20 | 0.04 | 0.01 | **Walking** | -0.50 | -0.29 |
|  | (0.12) | (0.09) | (0.02) |  | (0.43) | (0.45) |
| **Transit Stations** | 0.44 | 0.42 | 0.00 | **Driving** | -0.05 | 0.26 |
|  | (0.44) | (0.41) | (0.14) |  | (0.41) | (0.35) |
| **Work Places** | 0.81*** | 0.36* | 0.30* | **Transit** | 1.81*** | 0.99 |
|  | (0.22) | (0.19) | (0.16) |  | (0.53) | (0.64) |
| **Residential** | 2.39*** | 1.21* | 0.13 |  |  |  |
|  | (0.78) | (0.71) | (0.34) |  |  |  |
| **Observations** | 4799 | 4775 | 2599 | **Observations** | 1170 | 1071 |
| **Adjusted R-squared** | 0.657 | 0.735 | 0.889 | **Adjusted R-squared** | 0.780 | 0.782 |
| **Date Fixed Effects** | Global | Regional | U.S. | **Date Fixed Effects** | Global | Regional |

**Table 6:** Regression results relating estimated *R* values to 7-day lagged Google workplace mobility (top panel) and Apple driving mobility (bottom panel) indicators in the international panel. Columns 1-2: sample is restricted to before April 7th. Columns 3-4: after April 7th. All regressions include country fixed effects. Standard errors, clustered by country, are reported in parentheses. Stars indicate statistical significance (*p<0.1, **p<0.05, ***p<0.01).

|  | (1) | (2) | (3) | (4) |
| --- | --- | --- | --- | --- |
|  | Before April 7th |  | After April 7th |  |
|  | Google Workplace Mobility |  |  |  |
| Mobility (7-day lagged) | 0.89*** | 0.63*** | 0.23*** | 0.26*** |
|  | (0.21) | (0.19) | (0.08) | (0.08) |
| Observations | 1949 | 1925 | 2853 | 2853 |
| Adjusted R-squared | 0.658 | 0.719 | 0.801 | 0.821 |
|  | Apple Driving Mobility |  |  |  |
| Mobility (7-day lagged) | 1.07*** | 1.01*** | 0.12 | 0.25 |
|  | (0.20) | (0.27) | (0.17) | (0.18) |
| Observations | 1525 | 1488 | 1769 | 1740 |
| Adjusted R-squared | 0.711 | 0.728 | 0.895 | 0.901 |
| Date F.E. | Global | Regional | Global | Regional |

Table 1 and the left panel of Figure 4 provide results of regressions that use estimates of the effective reproduction number (*R*) from Kučinskas (2020) as the outcome variable, and a seven-day lagged measure of workplace mobility from Google data. Google mobility data is coded here as the fraction decline from baseline levels.^14^ The coefficients should therefore be interpreted as the associated decline in the effective reproduction number associated with a 100 percentage point (p.p) decline in mobility.

All specifications include country fixed effects (intercepts) to account for all cross-country differences in transmission stemming from time invariant country characteristics. To account for time trends, the specification in Column 1 includes global date fixed effects and the specification in Column 2 includes region-by-date fixed effects. The specification in Column 3 includes country-specific linear time trends. The specification in Column 4 flexibly controls for the number of days elapsed in each country since it confirmed its 100th Covid-19 case, using fixed effects for each value of this lapse.

Across the four specifications reported in Table 1, we estimate a positive and statistically significant relation between effective reproduction numbers, *R*, and Google mobility levels, indicating that increased mobility is associated with increased transmission. The coefficient on lagged mobility ranges from 0.72 to 0.53, depending on the specification. The estimates imply that a ten percentage point drop in the Google mobility measure is associated with a decline of between 0.05-0.07 units of *R*, depending on the specification used.

Figure 5 plots the country-level fixed effects estimated in specification (1) in Table 1. The figure provides a measure of the reduction in transmission rates that is unexplained by mobility levels. The figure also plots the fixed effects of a variant of specification (1) which uses region, as opposed to country, fixed effects. A clear ranking emerges in regions’ ability to reduce transmission rates using non-mobility suppression methods, with Asia most successful, North America least successful, and European countries in between.^15^

**Figure 5:**
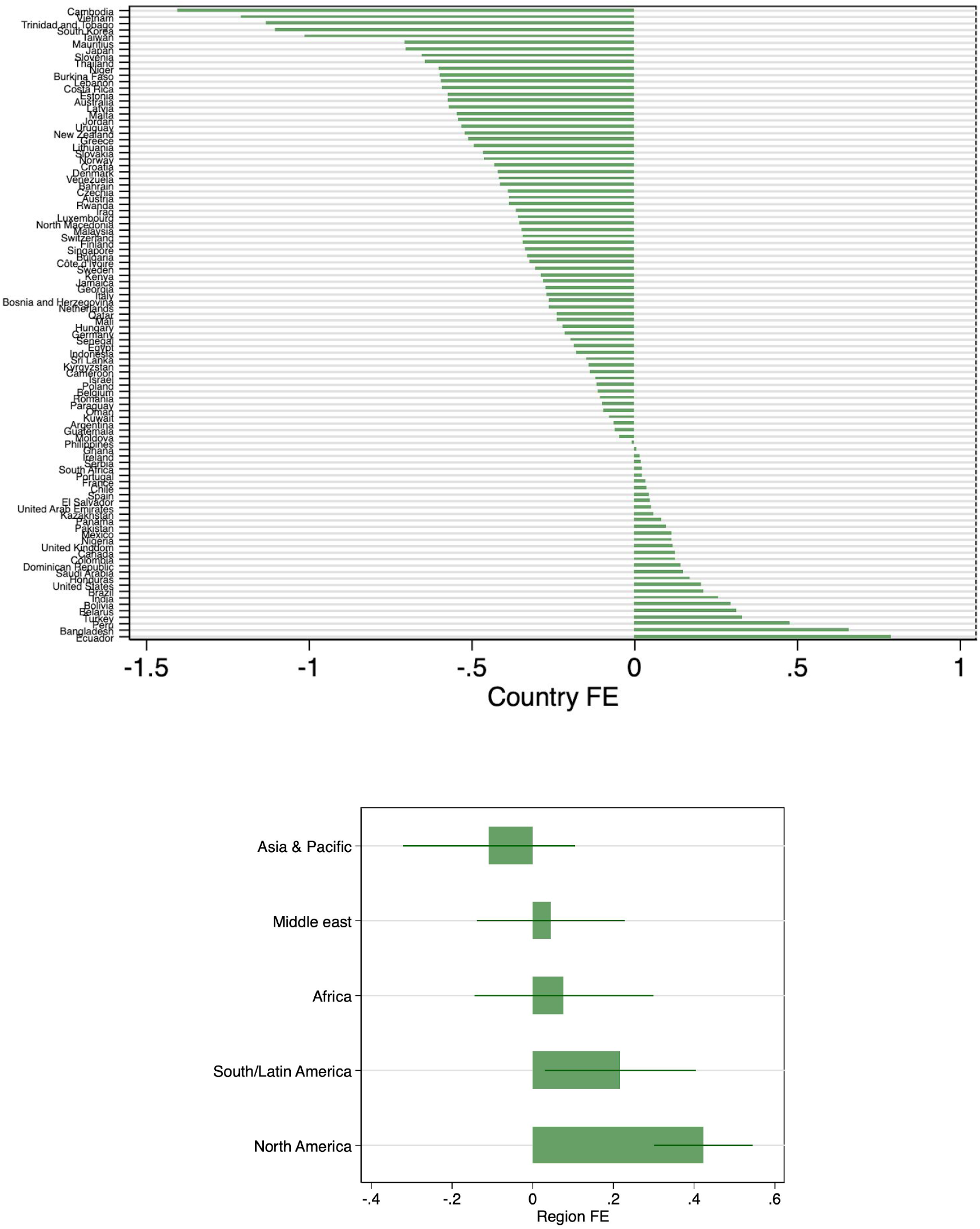
Top: Estimated country fixed effects from regression (1), sorted from smallest to largest by country. Bottom: region fixed effects from a version of regression (1) which replaces country fixed effects with region fixed effects.

Figure S6 plots the day fixed effects estimated in specification (1), which reflect the average daily variation in transmission that is unexplained by mobility reductions or by time-invariant country specific factors. As can be seen, the day fixed effects decline over time, indicating increased global suppression of transmission rates stemming from measures unrelated to variation in mobility (these measures could potentially include increased usage of masks, increased hygiene, favorable weather trends, etc).

In a secondary analysis (Table S1), we also use related, but somewhat cruder indicators of transmission as the outcome variables, which consist of the growth rates of the total number of cases and deaths and the growth rates of the daily new cases or deaths. The results are consistent with our primary specification.

The right panel of Figure 4 and Table 2 repeats the analysis in Table 1 but uses the Apple driving mobility measure to proxy for mobility levels. Results are similar to those in Table 1 and indicate positive and statistically significant relations between *R* and mobility in all four specifications. The estimates imply that a ten percentage point drop in the Apple mobility measure is associated with a decline of 0.07–0.08 units of *R*, depending on the specification used.

Table 3 examines the relation between transmission rates and mobility levels in an analogous manner, but uses the state-level panel dataset in the United States. Transmission rates are taken from Systrom and Vladeck (2020), and mobility is measured either through Google workplace mobility or Apple driving mobility indicators. All regressions include state fixed effects and either date fixed effects or state-level time trends. As can be seen, similarly to the global analysis, the results in the United States indicate a positive relation between transmission rates and mobility levels. Focusing on Column 1, which is most similar to our benchmark global model (i.e., with country and date fixed effects), the estimates suggest that a 10 percentage point reduction in mobility levels is associated with a 0.04 decrease in the value of *R*. The size of the effect is rather similar to that in the benchmark global estimate (0.06) despite the different geographic context and the fact that the estimates of *R*(*t*) used in the two regressions are derived by independent methods and researchers.

The top panel of Table 4 repeats the global estimation model separately in five different geographical regions using Google mobility data (see also Figure 4).^16^ As can be seen, the positive relation between transmission rates and mobility levels is concentrated in Europe, the “Asia and Pacific” region, and the South/Latin America region. No statistically significant relation between transmission rates and lagged mobility is observed in the Middle East or Africa, perhaps owing to the relatively small number of observations within these regions.^17^ Employing the Apple driving mobility measure (bottom panel in table), shows that the relation between transmission rates and mobility is concentrated in countries within Europe, with imprecision for other regions potentially stemming from the substantially lower country coverage offered by the Apple mobility data and the resulting sample size.

We next conduct an exploratory analysis of how infection rates relate to the various mobility measures provided by Google and Apple. Results should be interpreted with caution given the high levels of temporal correlation between the various location-based Google mobility measures (workplace, residential, parks, retail and recreation, transit, grocery and pharmacy) as well as the high correlation between the search-type Apple mobility measures (driving, transit, walking). Table 5 reports estimates of regressions (1) and (2), respectively, that include as independent variables all six location-based measures of Google mobility (with date and date-by-region fixed effects, respectively). Columns 3 and 4 report the analogous specifications that simultaneously include all three search-type Apple measures. Figure 6 compares the coefficients on the various measures of Google mobility when estimated separately and simultaneously.

**Figure 6:**
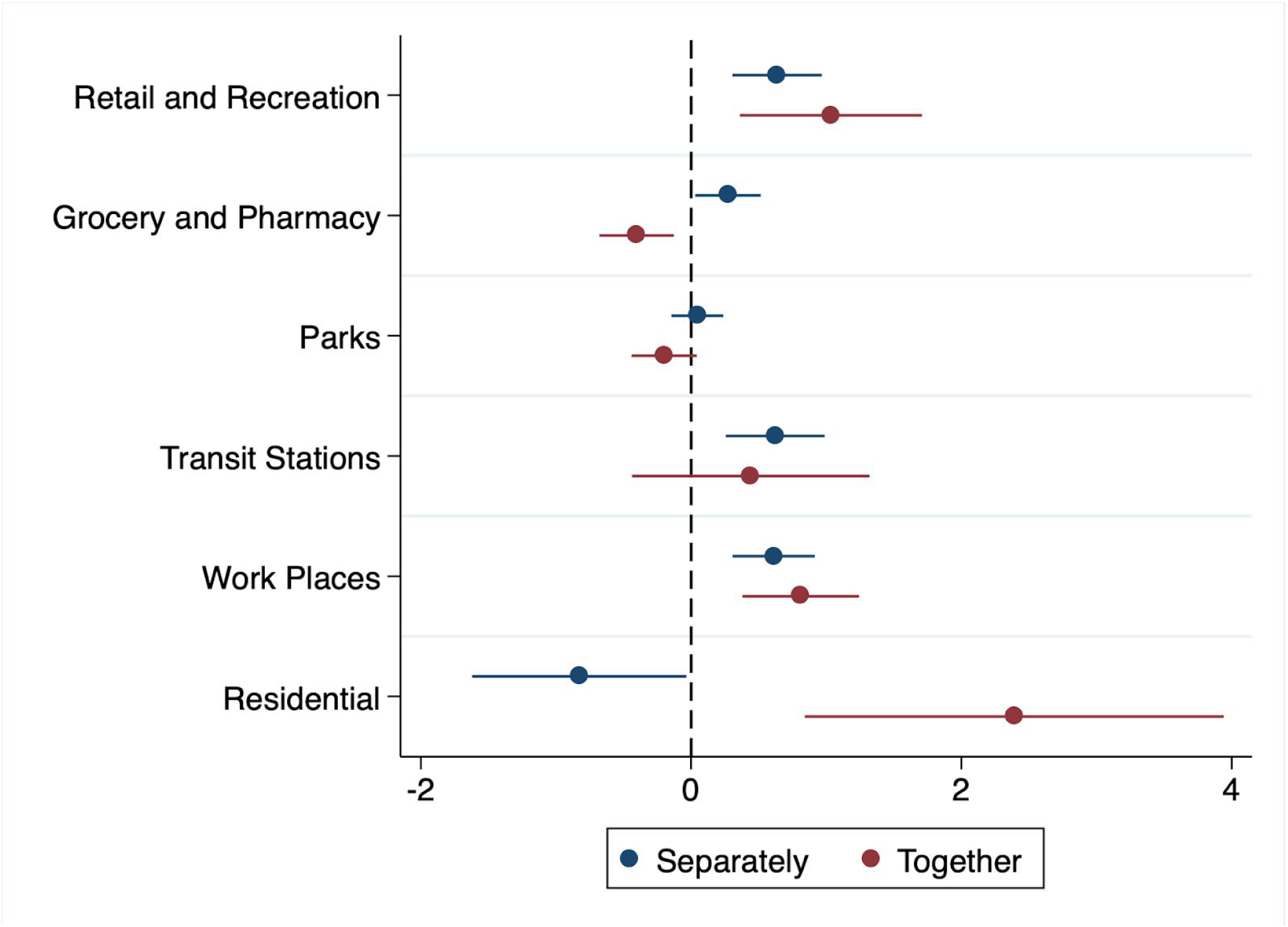
Coefficients from regressions of estimated effective reproduction numbers *R*(*t*) observed at the country-date level on six Google workplace mobility indicators, estimated in separate regressions (blue markers) or together in a single regression (red markers). Markers indicate point estimates and error bars indicate 95% confidence intervals corresponding to standard errors that are clustered at the country level. The model includes country and date fixed effects.

Focusing on the first column of Table 5, the results provide suggestive evidence that increased mobility in “work places”, “retail and recreation”, and “residential” areas are positively associated with transmission rates. Coefficients on the Google location-based measure should be interpreted in relation to an omitted location category defined as the complement to the union of the six locations defined by Google. Similarly, the results in Column 1 indicate a negative relation between transmission rates and increased population mobility in “grocery and pharmacy”. These results continue to hold after adding regional date fixed effects (Column 2) but the magintudes of the coefficients are reduced. The positive coefficient on “Residential Areas” is somewhat counterintuitive. One potential explanation might stem from increased prevalence of infection between members of the same household (see, e.g., Bi et al., 2020; Qian et al, 2020). Regional inspection suggests this effect is concentrated in Asia (results not shown).

Column 3 provides a parallel analysis to that reported in Column 2, but uses the U.S. state panel with estimates of infection reproduction numbers taken from Systrom and Vladeck, 2020. Similar to Column 2, results indicate that workplace mobility in the United States is positively related to transmission rates in a statistically significant manner, with the coefficient on mobility estimated at 0.36. All other mobility measures in Column 3 are non-significant.

Columns 4 and 5 of Table 5 use Apple search-type mobility data. Focusing on Column 4, when all three mobility measures are included in the regression, the coefficient on transit related searches is the only measure which is statistically significant, while the other mobility measures based on driving and walking—activities involving less social interactions—are not related to *R* in a statistically significant manner. However, once regional fixed effects are added, none of the coefficients associated with the three Apple mobility measures are statistically significant, potentially due to their high levels of correlation. As additional data becomes available, we will continue to update these estimates with the hope that increased sample size may allow for greater precision.

We continue by analyzing evidence for a structural break in the relation between transmission rates and mobility levels during our sample period. As shown in Figure 2, countries in Europe and the Asia and Pacific region exhibit an initial large decline in both Google workplace mobility as well as transmission levels up to late March. This is followed by a phase of relative stabilization in workplace mobility but a continued decline in estimated reproduction numbers. In some countries, there are indications of rising mobility levels towards the end of April. We therefore examine the possibility that the different nature of the “lockdown” and later phases can result in differences in the strength of the mobility-transmission relation.

Table 6 repeats the baseline analysis reported in Table 1, but separately conducted over the time period up to, and following April 7th (because of the 7-day lag, this means the data used in these two analyses extends up to and after April 1st). Results in the table provide suggestive evidence for a weakening in the relation between transmission rates and mobility, with larger point estimates and higher significance levels in the three specifications estimated over the former part of the sample period as compared to those in the latter part of the sample period. Focusing on the top panel (using Google workplace mobility measures) and comparing columns (1) and (3) shows that the relation between transmission rates and mobility estimated after April 7th is roughly 25% of that estimated prior to April 7th. Using the Apple driving mobility measure (bottom panel), we do not find a statistically significant relation between transmission rates and mobility levels after April 7th.

The decline in the relation between transmission rates and mobility levels could be explained by policy responses undertaken at the country-level or individual-level behavioral changes. Additionally, environmental changes could also weaken the association between mobility and Covid-19 transmission. We note that mobility levels in a number of countries have begun rising in the latter part of the sample period. As transmission rate indicators arrive with a lag, additional data arriving in the upcoming weeks will be of particular interest in tracking the evolution of mobility levels and Covid-19 transmission rates.

## Discussion

Given the estimates in Table 1, a 10 percentage point decline in the Google workplace mobility measure is associated with a 0.05–0.07 unit decline in the estimated reproduction number *R*. Based on these estimates, it is instructive to analyze the share of the overall decline in transmission rates that can be explained by mobility reductions.

Overall, Google mobility rates in Europe declined by approximately 50 percentage points by the beginning of April, which our estimates imply is associated with a reduction in *R* of 0.25–0.35 units. The fraction of the overall decline in *R* that is explained by this mobility reduction depends on the time window chosen for the calculation. By April 11th, the mean value of *R* across the European countries in our sample had declined to 0.95.^18^ Taking the starting date for the calculation as February 21st, when *R* = 3.1, implies an overall decline in *R* of 2.2 units, meaning that mobility reductions explain about 12%-16% of this decline. However, data from February is still very sparse, and derived from only a few countries. If we more conservatively choose the starting time of the calculation to be a month later, on March 11th, when *R* = 2.1—at this point data is available from more than 20 countries, and most of the European mobility reduction has yet to take place—the overall reduction in *R* is reduced to 1.1 units, so that mobility reductions explain approximately 22%–30% of the decline in *R*.

Analogously, in countries within the “Asia and Pacific” region the decline in average Google mobility measures—approximately 40 percentage points to date—imply a reduction of 0.24–0.36 units in *R*. Again, the fraction of the overall decline in *R* explained by mobility depends on the time frame chosen. Taking the starting date of the calculation to be March 11 (average *R* = 1.7), and given the April 11th average value of *R* of approximately 1.27, we obtain that mobility reductions in Asia explain approximately 30%-41% of the reduction in *R*.

Our estimates, to a large extent, are estimated during a period when both mobility levels and transmission rates decline. Assuming that the association between mobility and *R* remains similar when the two variables rise, the same coefficients, interpreted at face value, also imply that in Europe, returning from current levels of mobility to pre-pandemic baseline levels—approximately a 50 percentage point increase—would entail an increase of between 0.25 (S.E. 0.08) to 0.35 (S.E. 0.08) points in *R*. Under this scenario, in order to avoid levels of *R*(*t*) from systematically climbing above the critical value of *R* = 1, easing of mobility restrictions may have to coincide with other *non-mobility* measures which reduce *R* to a level of 0.65–0.75.

Similarly, the coefficients imply that in Asia, returning to baseline levels of mobility—approximately a 40 percentage point increase—would entail an increase of between 0.20 (S.E. 0.06) to 0.28 (S.E. 0.06) points in *R*. To avoid levels of *R*(*t*) from systematically climbing above *R* = 1, easing of mobility restrictions in Asia may have to coincide with other non-mobility measures which reduce *R* to a level of 0.72–0.8.

Turning to the United States, the estimates based on the state-level analysis imply that a 10 percentage point decline in the Google workplace mobility measure is associated with a reduction of between 0.04 units (S.E. 0.013) and 0.05 units (S.E. 0.004) in the value of *R*. As during the sample period average reproduction numbers in the United States fell by approximately 0.5 units, this estimate suggests that the decline in mobility in the United States can explain approximately 32%-42% of the overall decline in *R*. This estimate, obtained using within country variation and a different source and methodology for estimating *R*, is rather similar to that obtained in Europe and Asia. Taken at face value, our estimates suggest that to avoid levels of *R*(*t*) from systematically climbing above *R* = 1, returning to pre-pandemic levels of mobility in the United States may have to coincide with other non-mobility measures which reduce *R* to a level of approximately 0.8.

Understanding the determinants of Covid-19 transmission rates is one of the most pressing policy questions facing society. This paper provides an empirical analysis of this question, utilizing comprehensive data at global scale to analyze the correlation between mobility levels and transmission rates over time within countries. As such, it provides an important complement to detailed epidemiological modelling of the spread of Covid-19. It will be valuable to revisit the estimates provided in this paper as mobility levels and Covid-19 transmission rates continue to vary in the coming months and as additional data become available.

## Data Availability

All data is publicly available as explained in the paper.

## Supplementary Information

**Figure S1:**
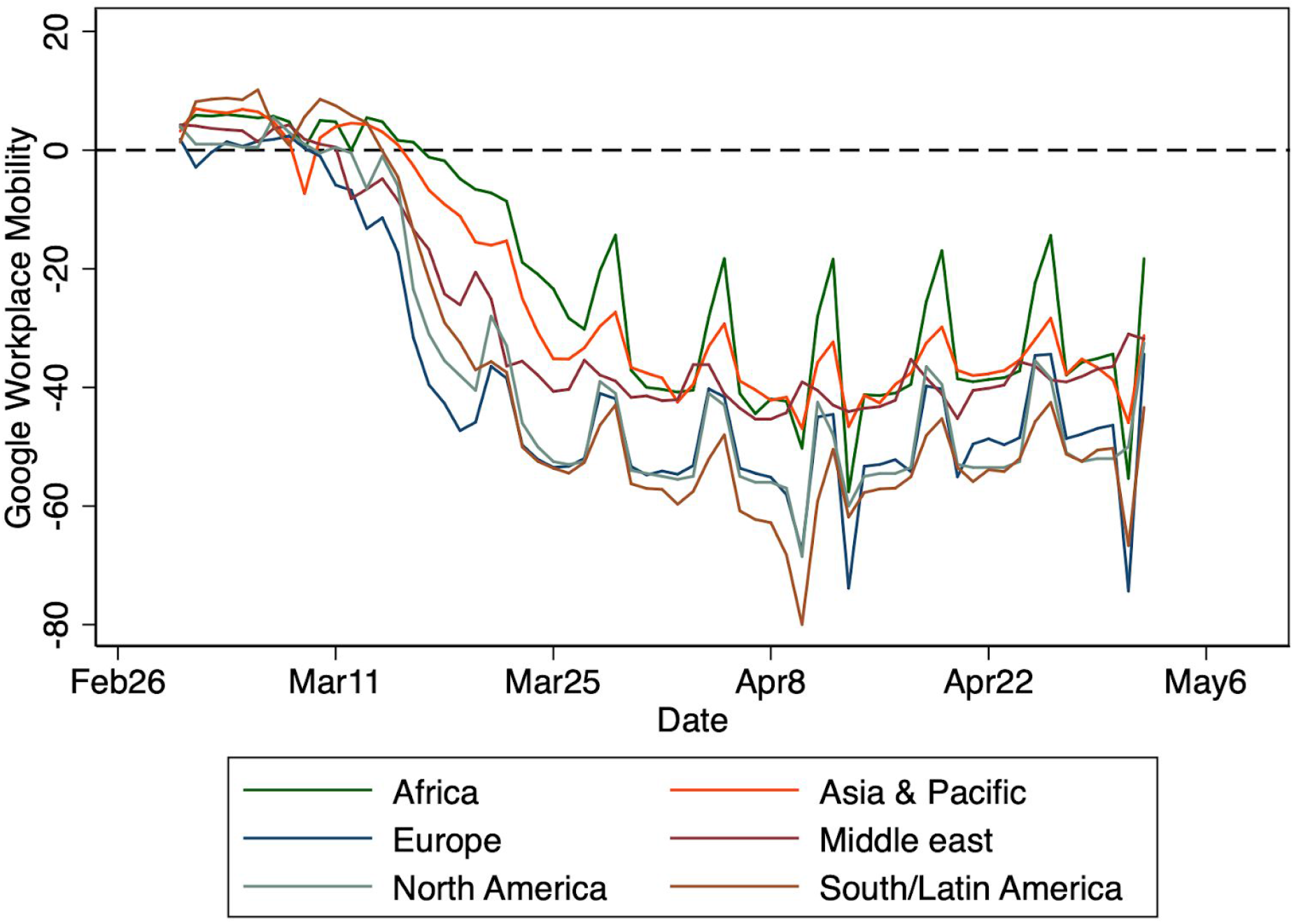
Average measure of Google workplace mobility over time, by region.

**Figure S2:**
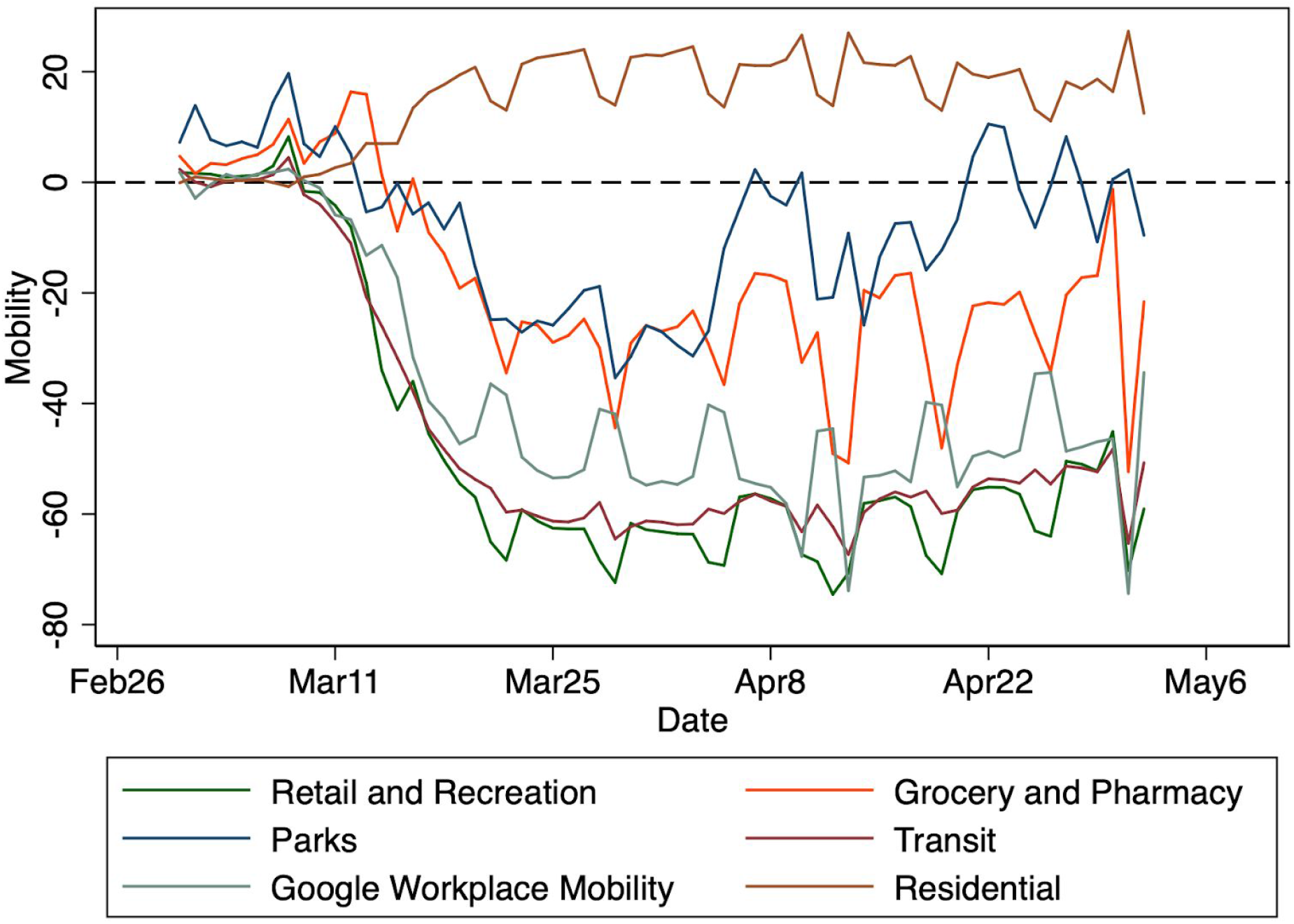
Average measures in Europe in Europe of Google mobility data, by mobility indicator type, by mobility indicator type, over time.

**Figure S3:**
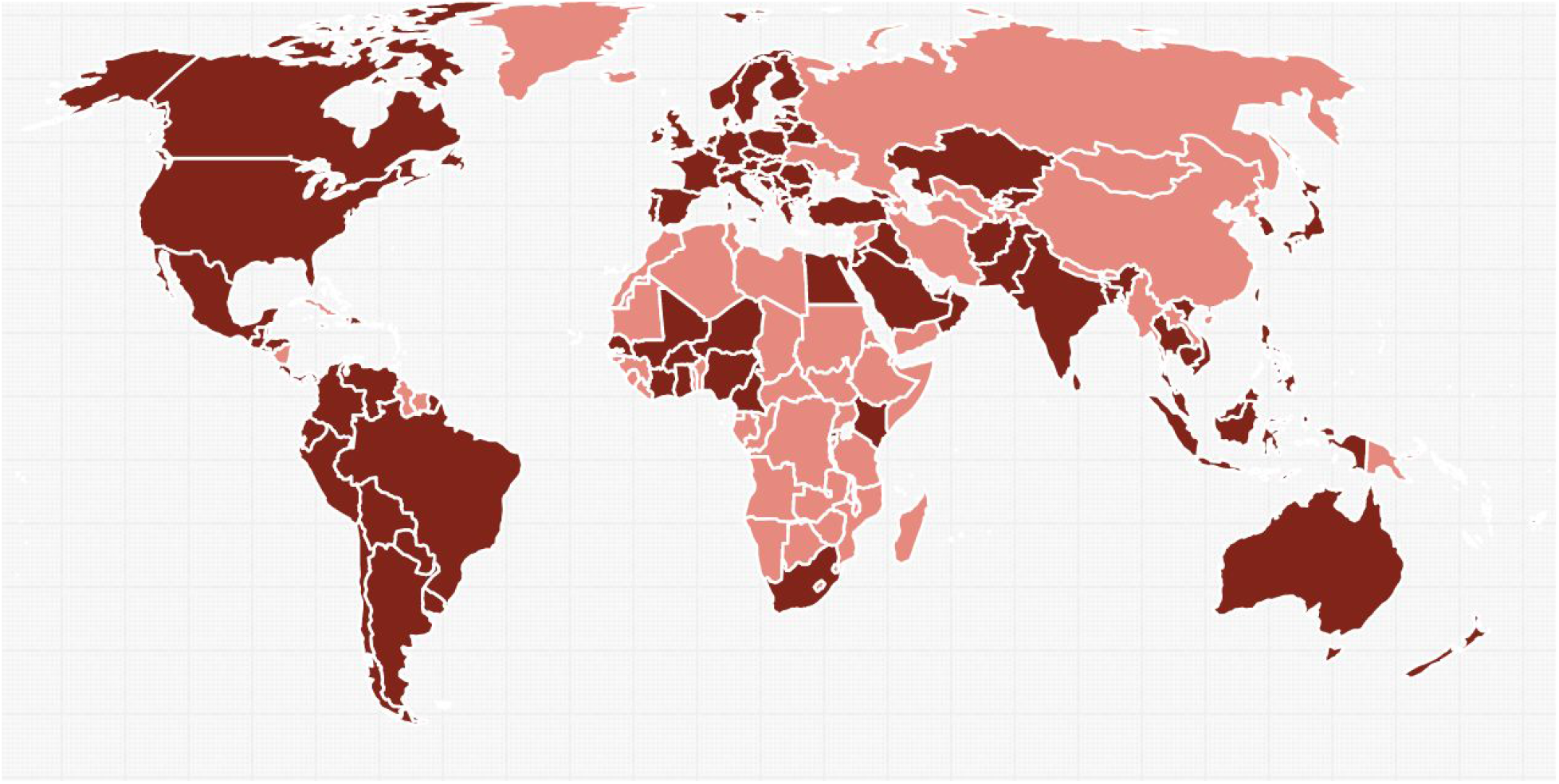
Countries included in the international sample (shaded in dark red). Prepared through simplemaps.com.

**Figure S4:**
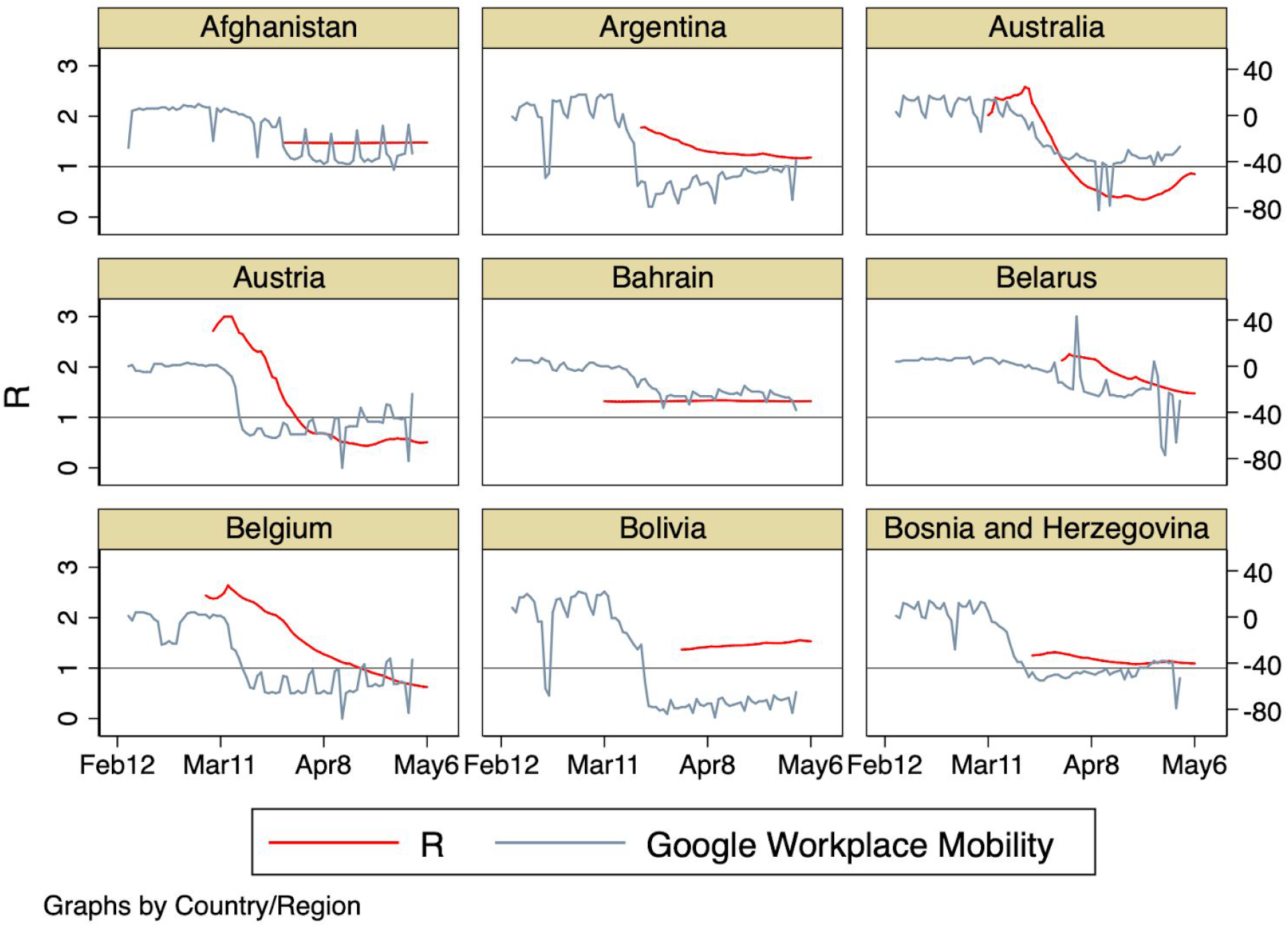
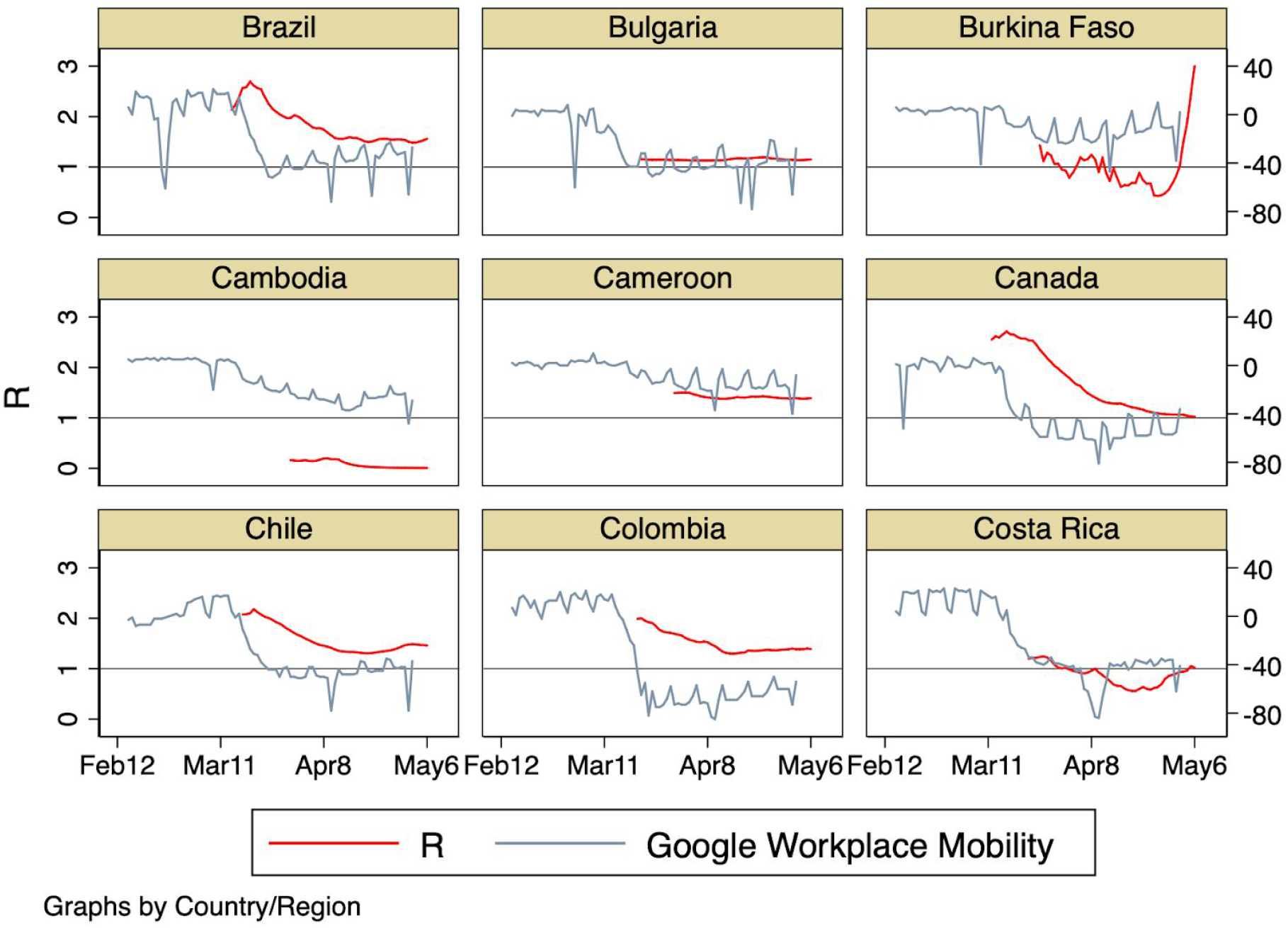
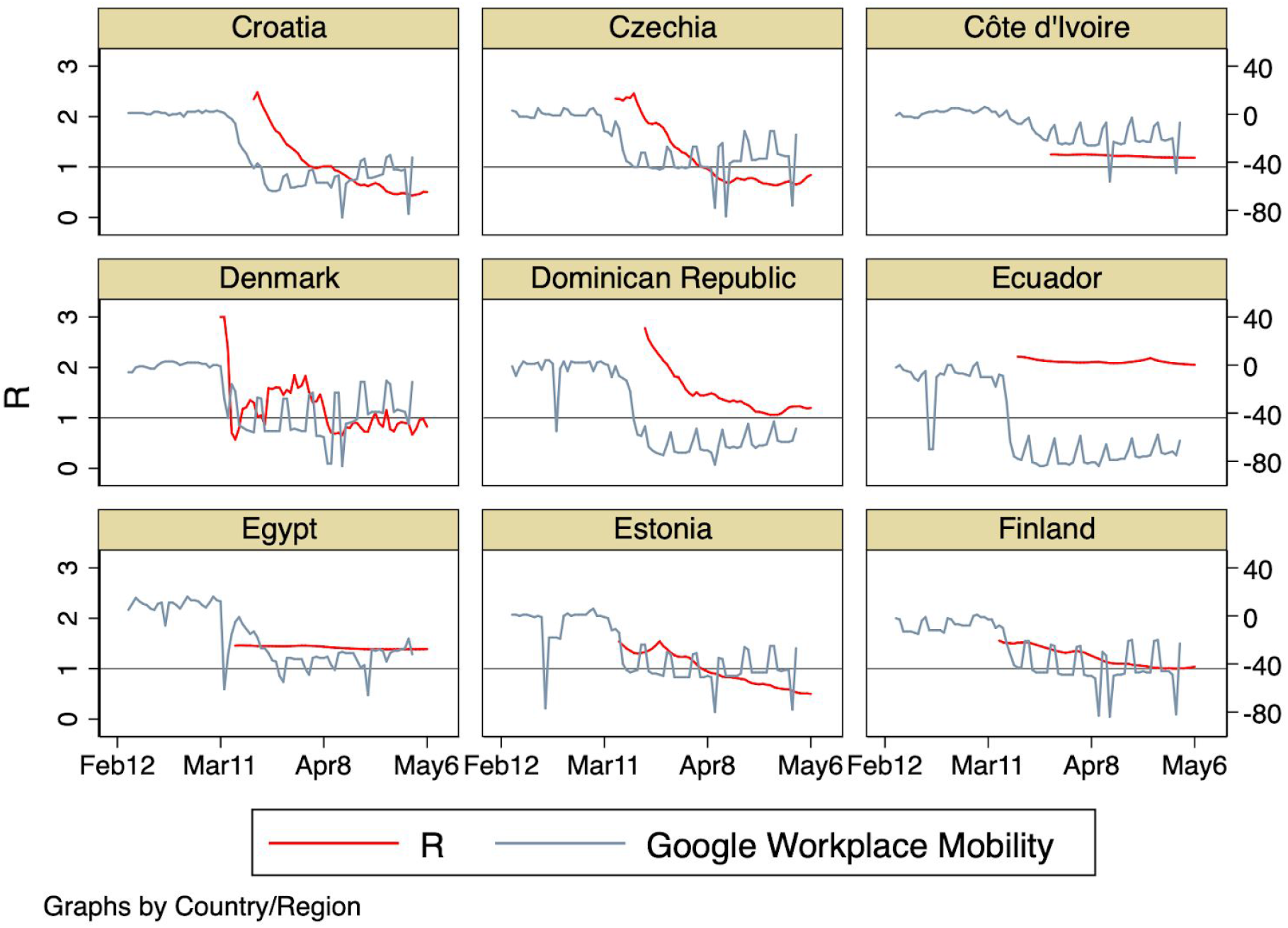
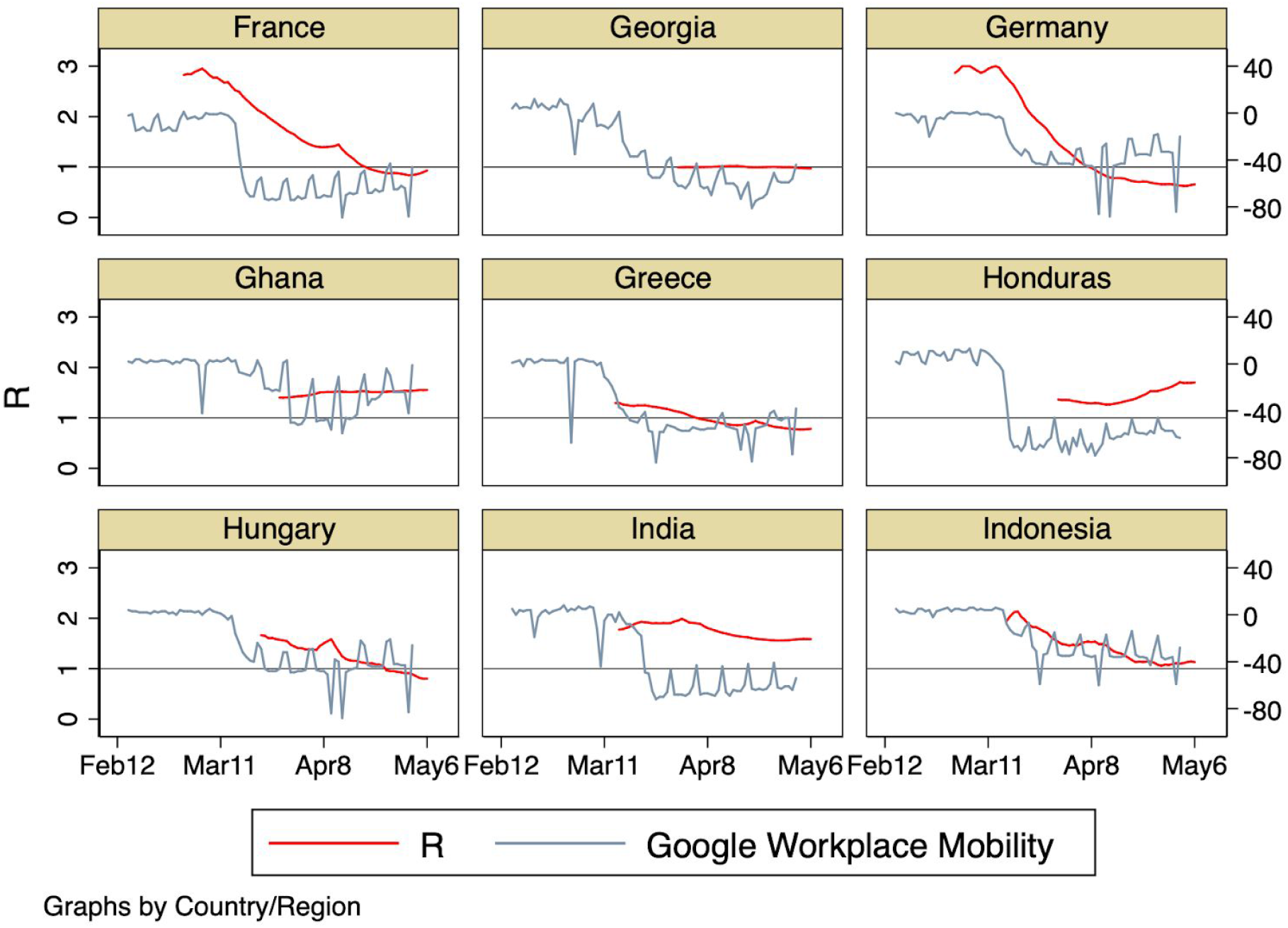
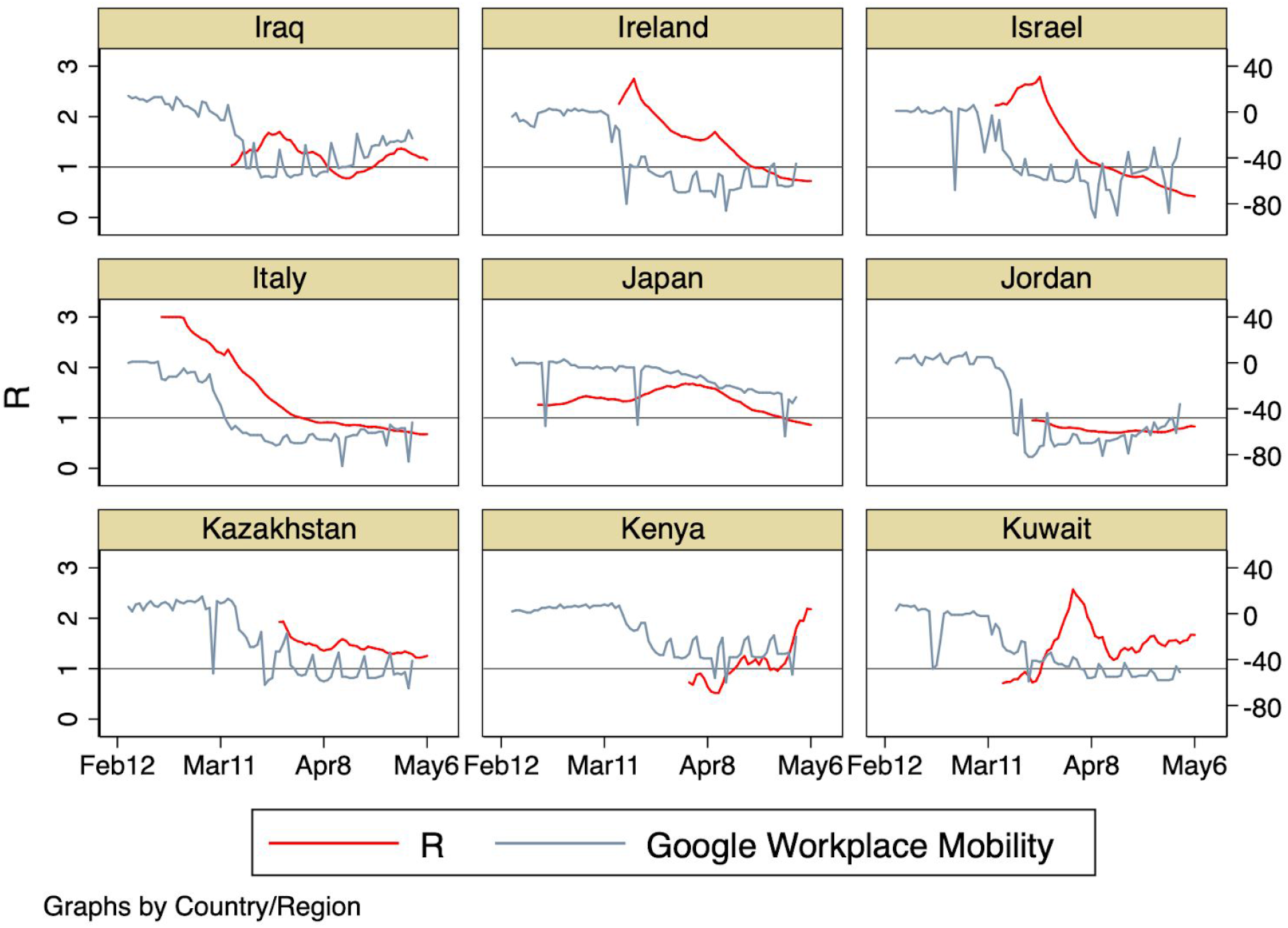
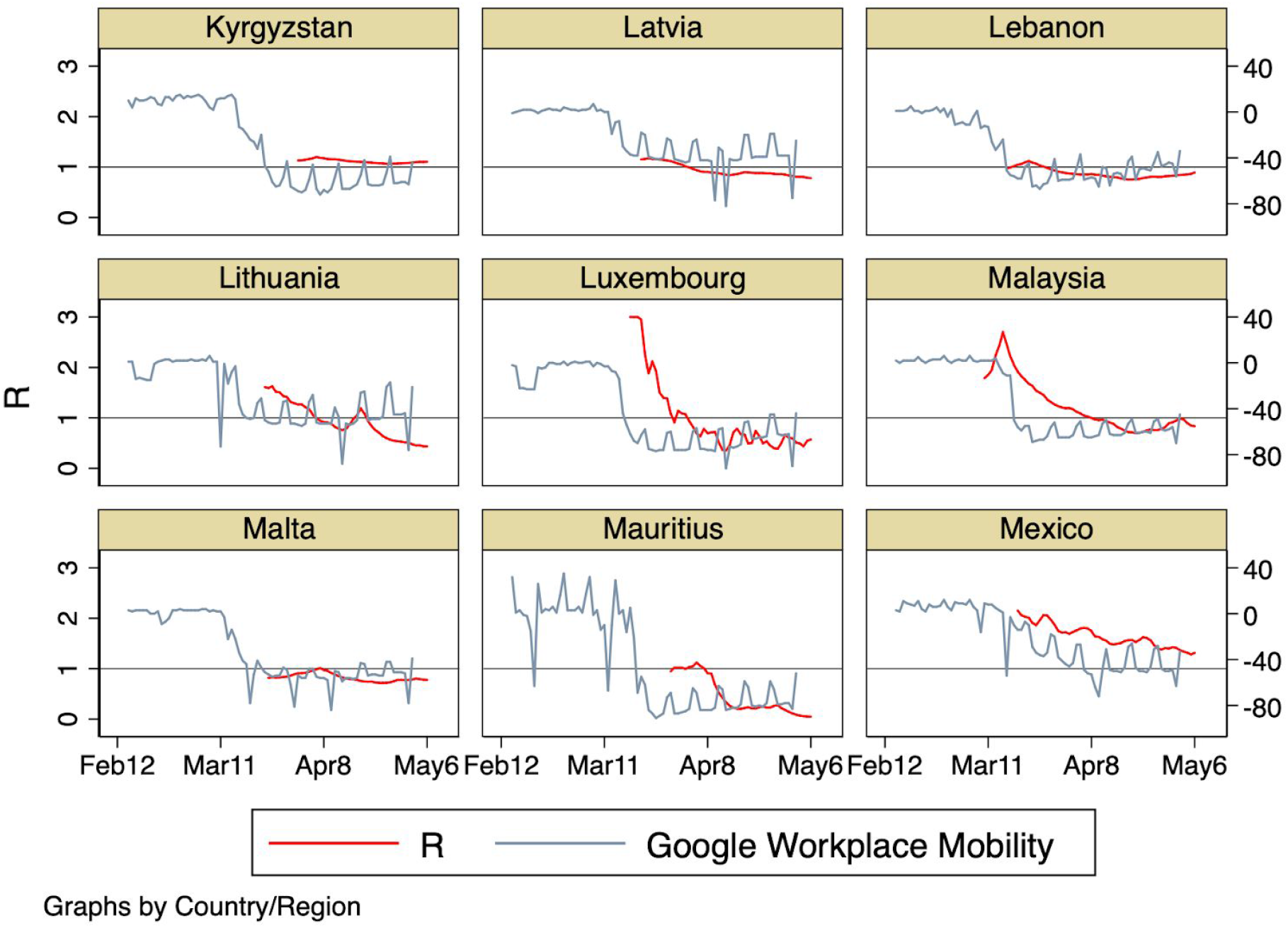
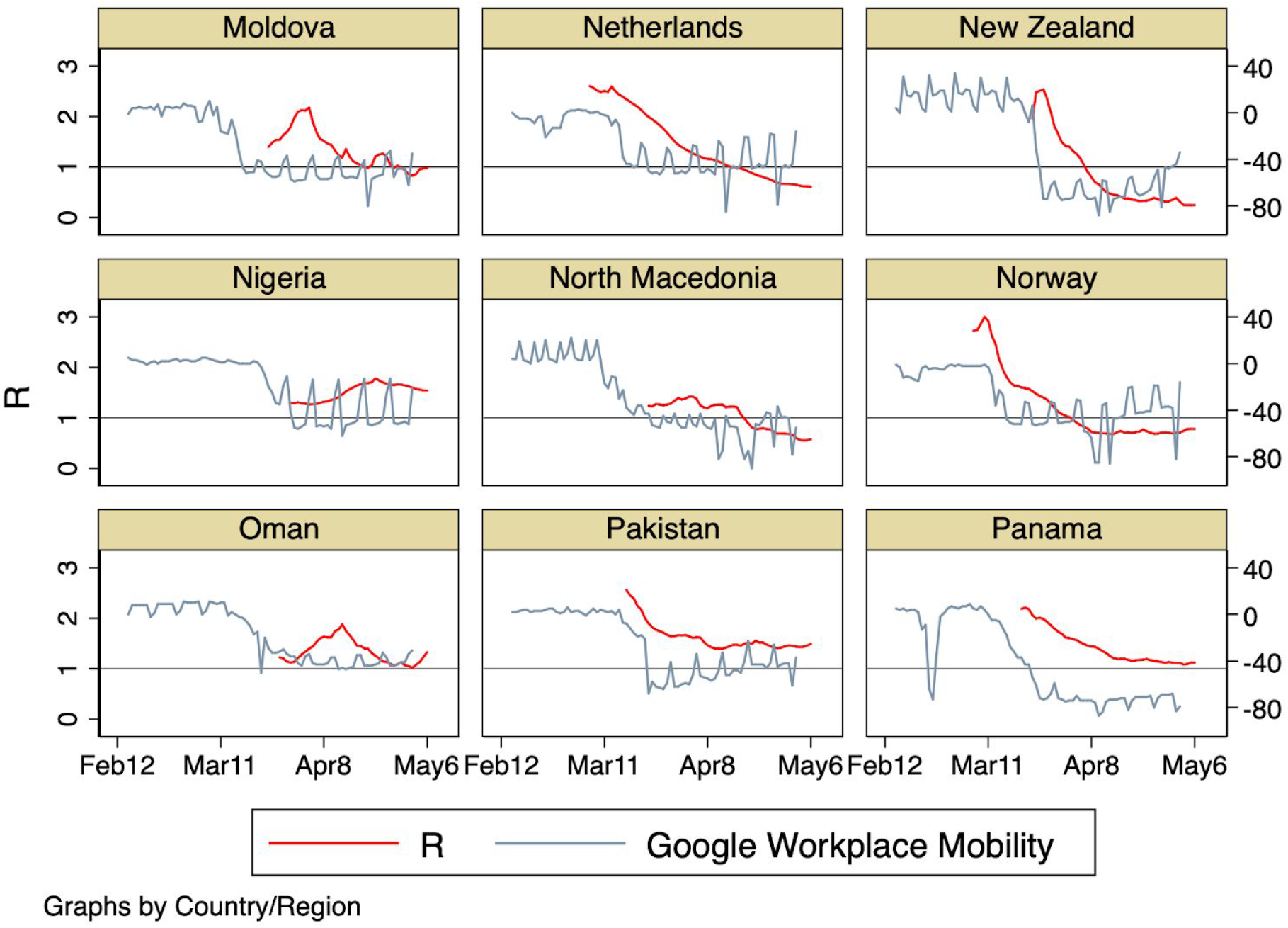
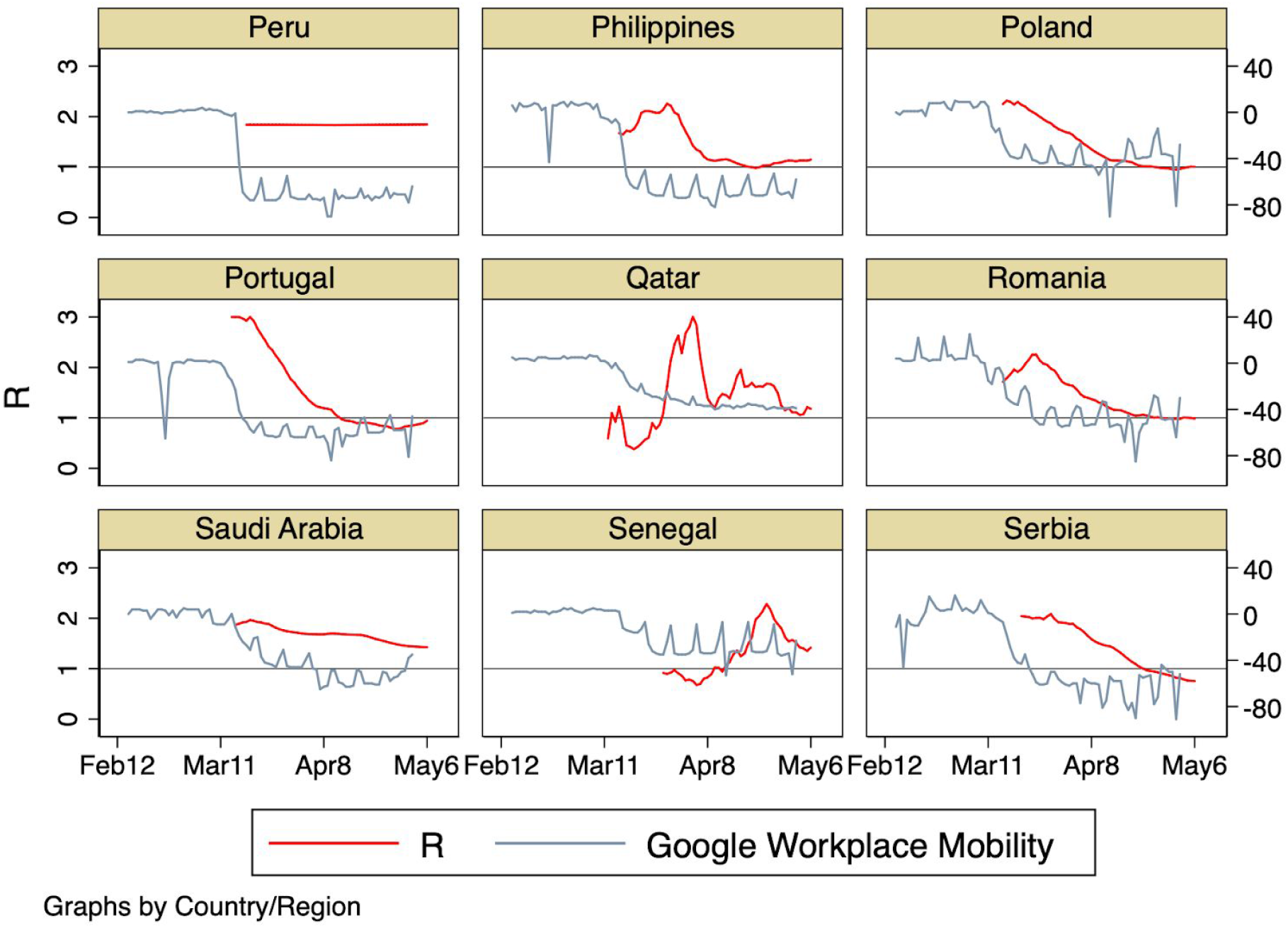
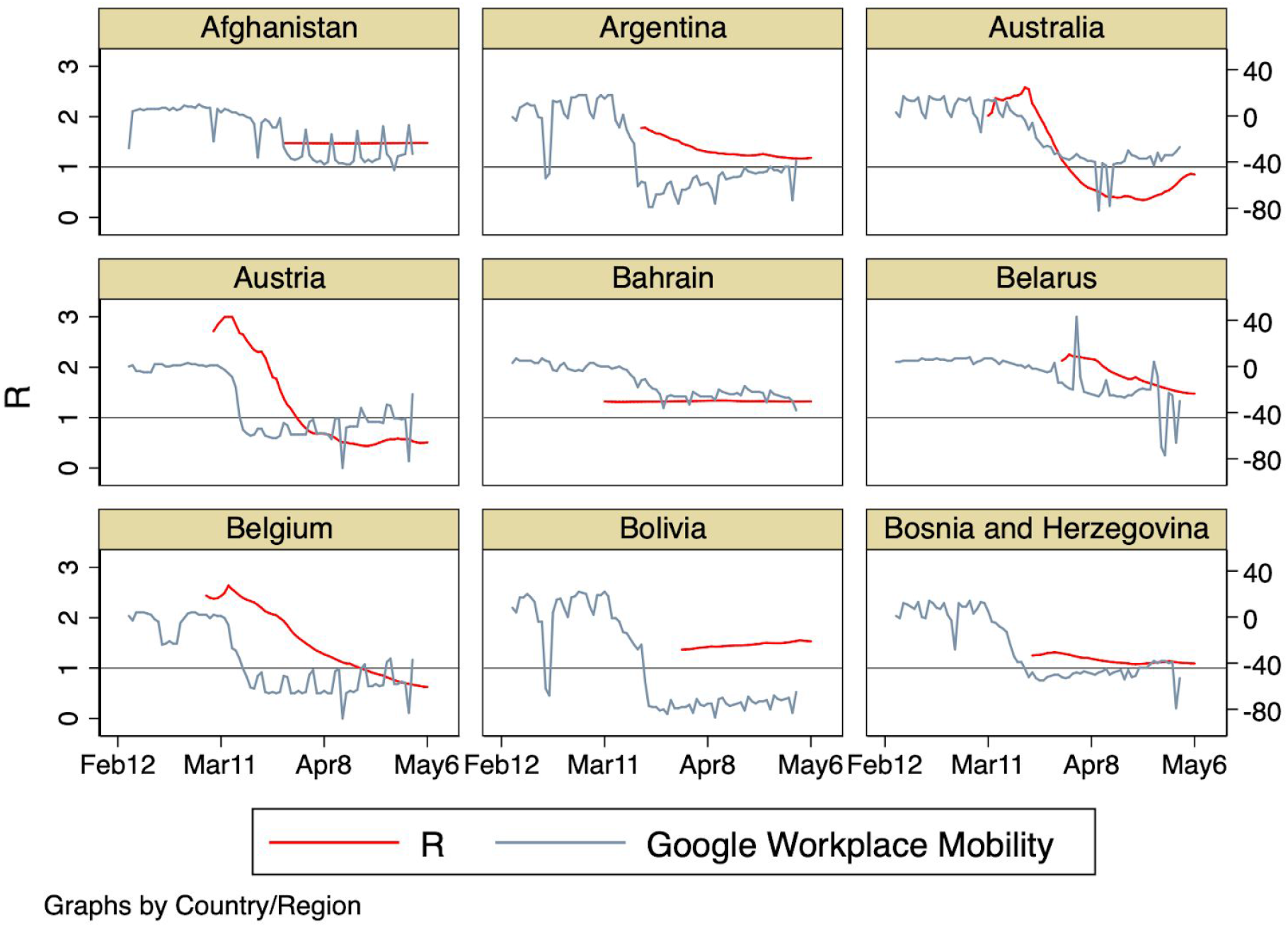
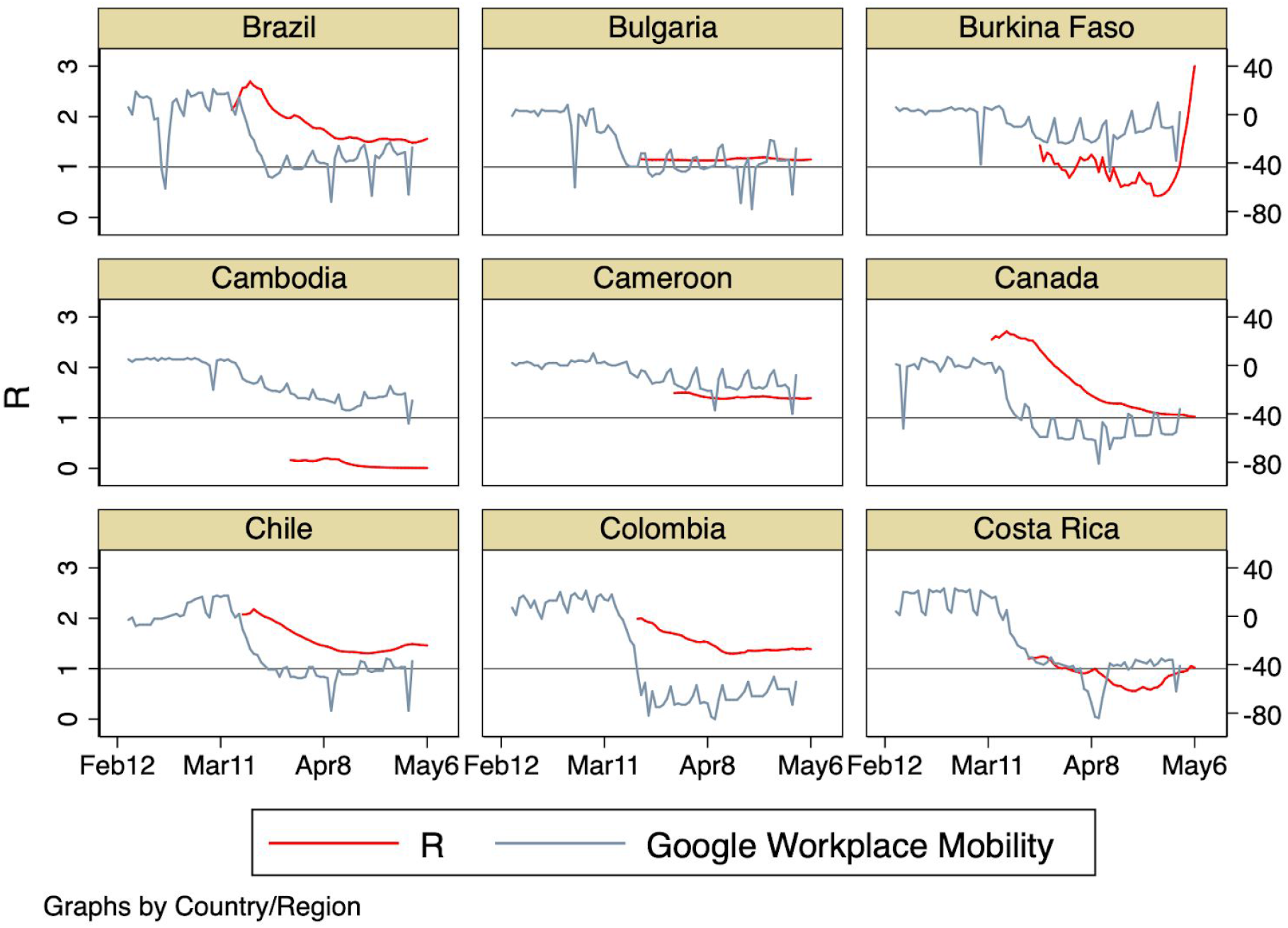
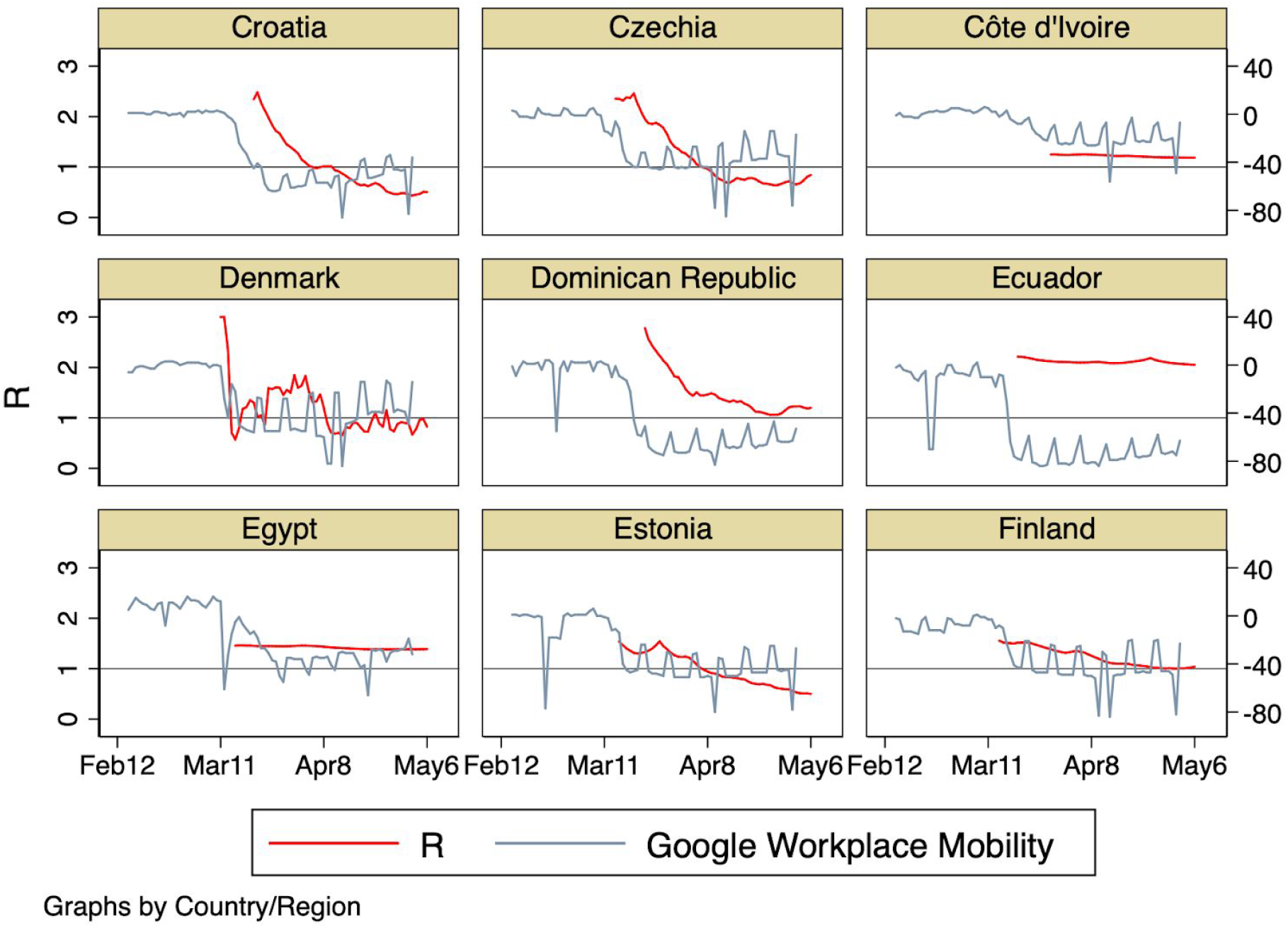
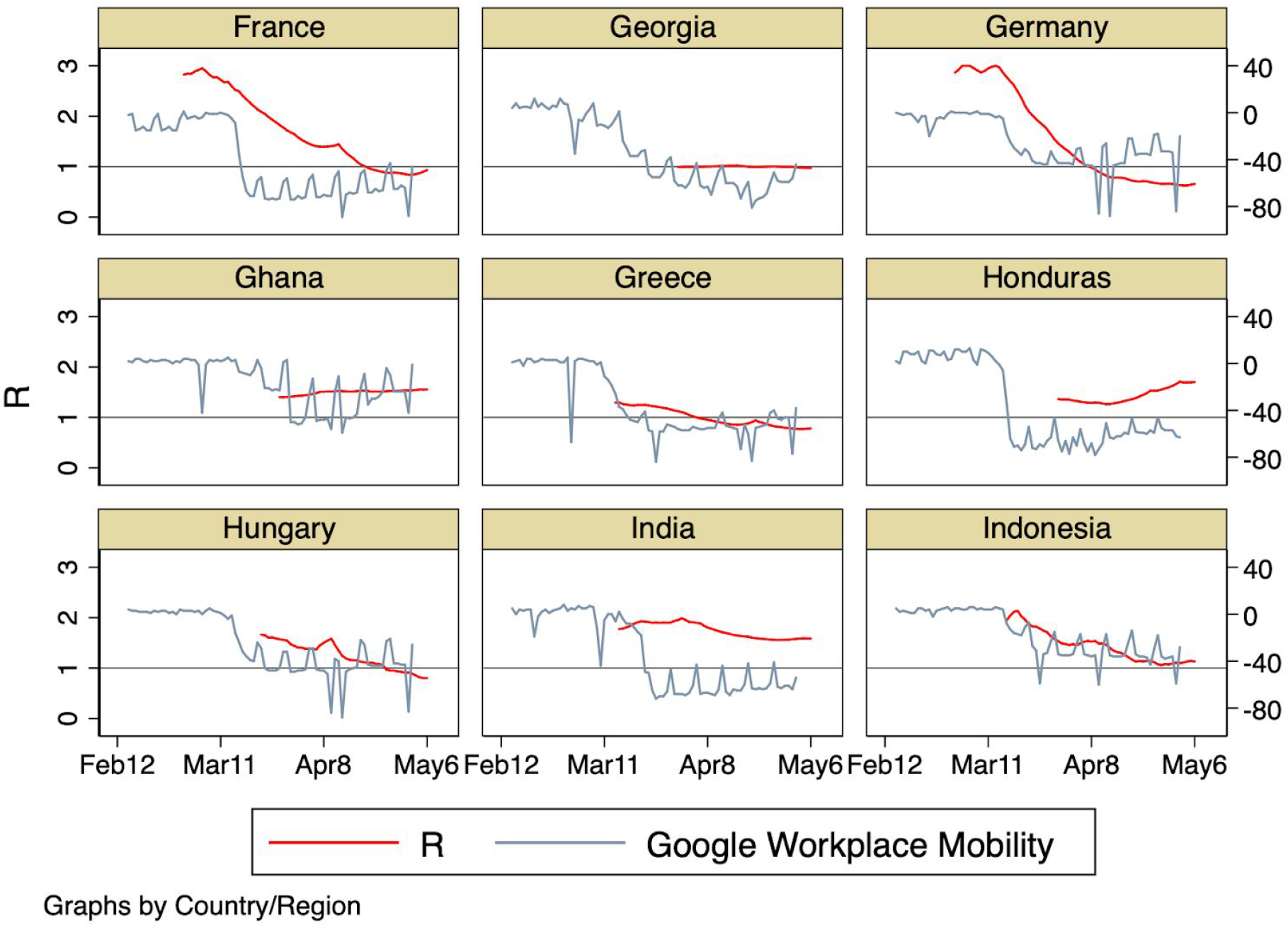
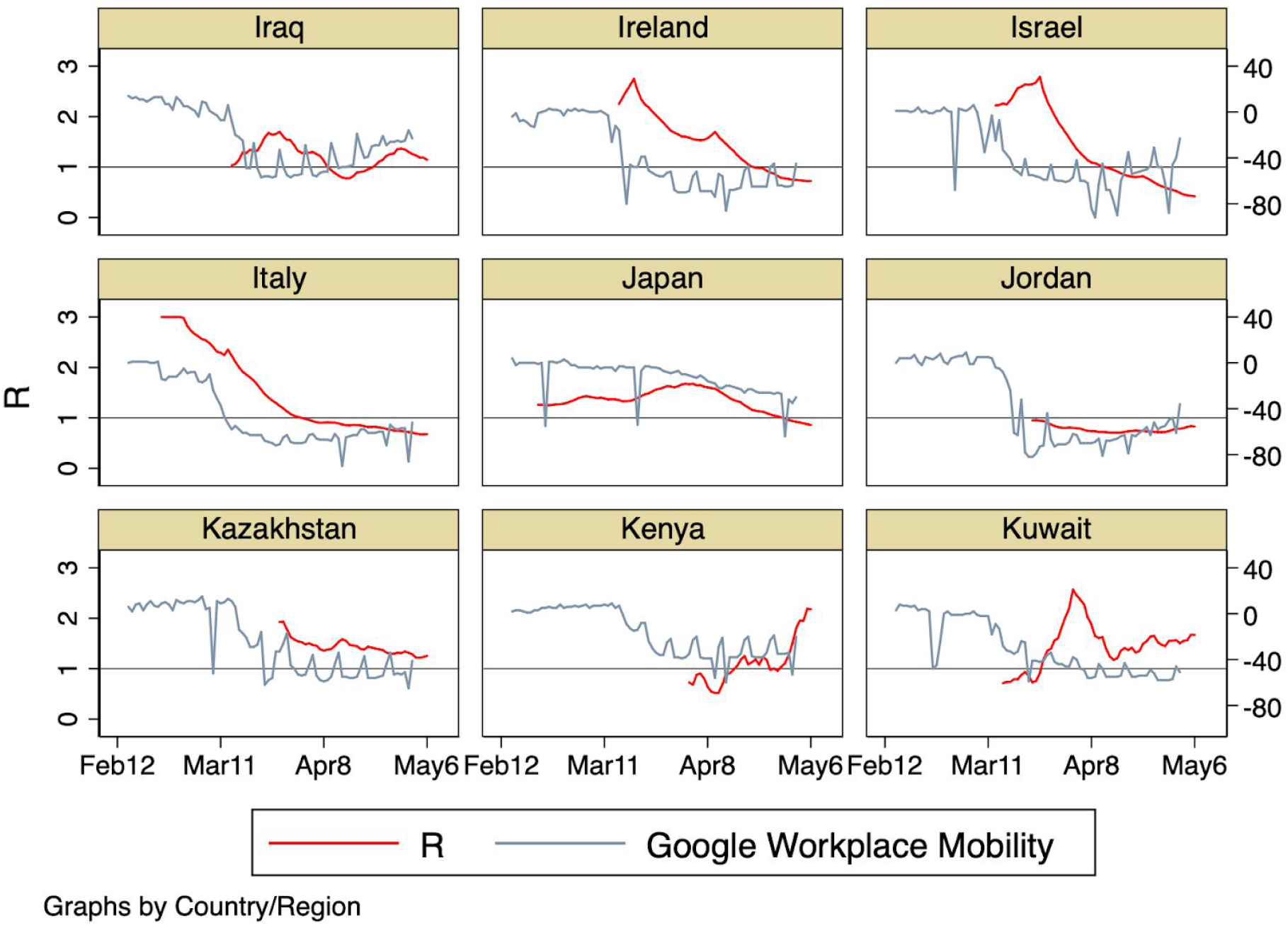
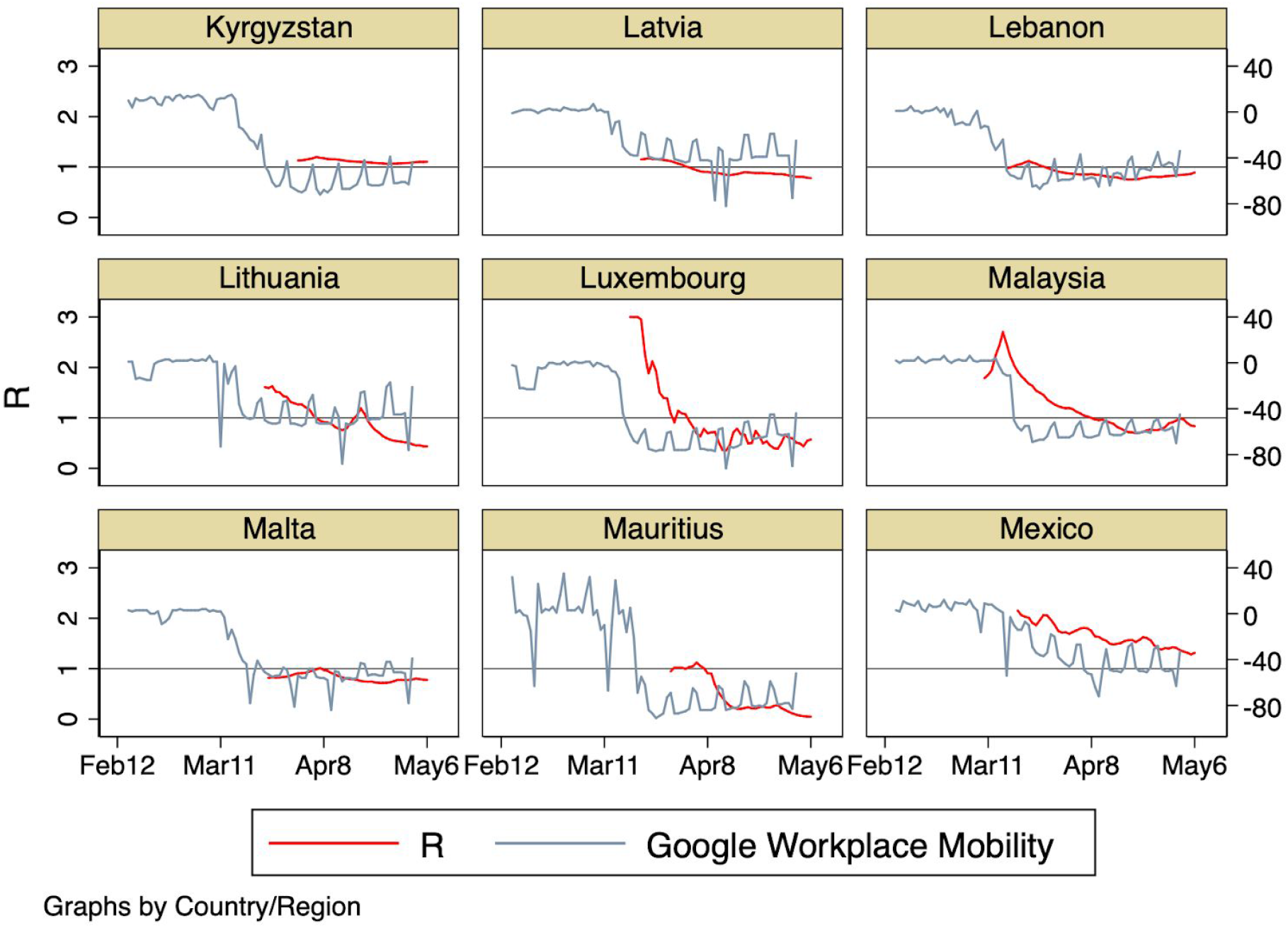
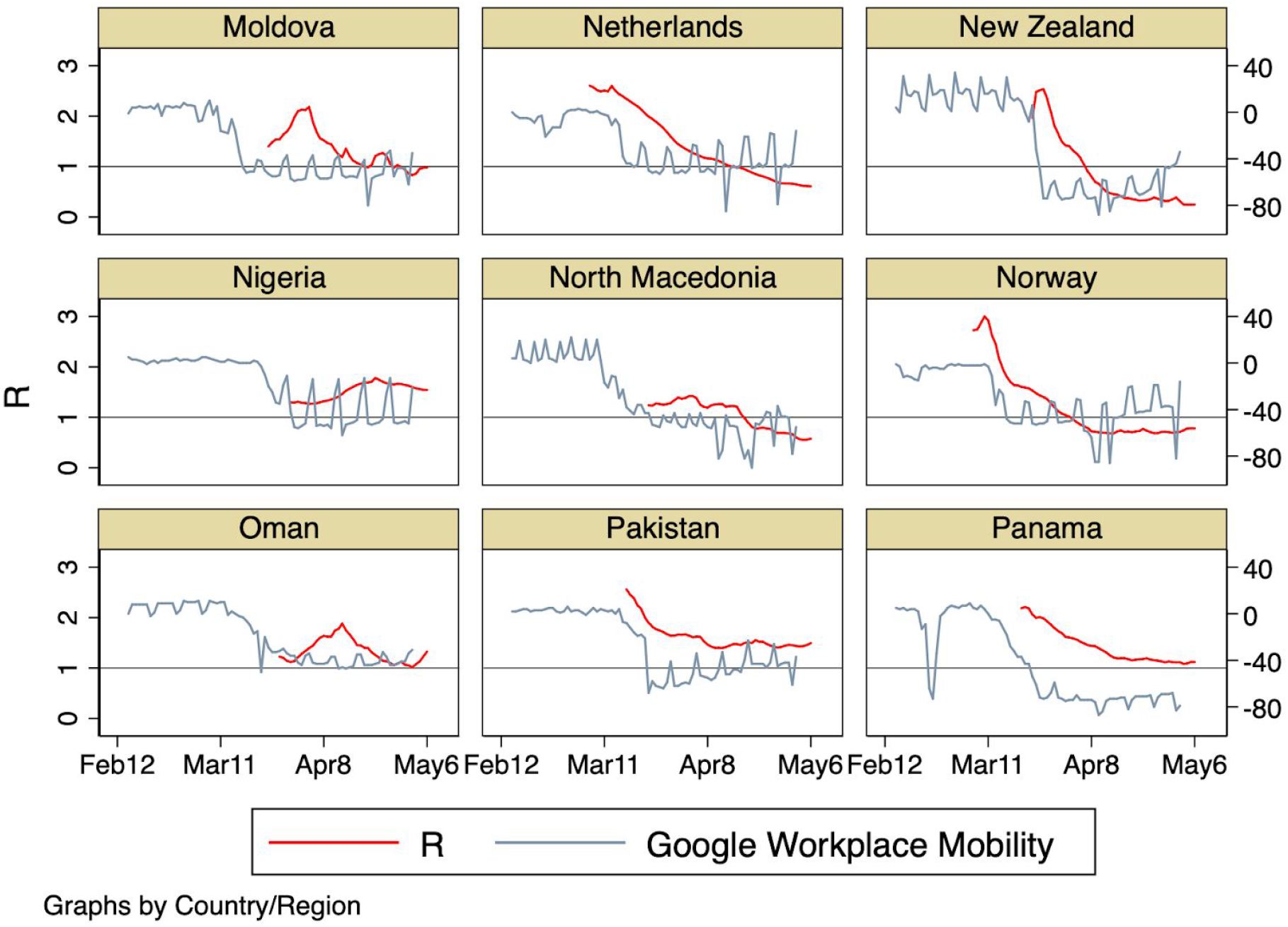
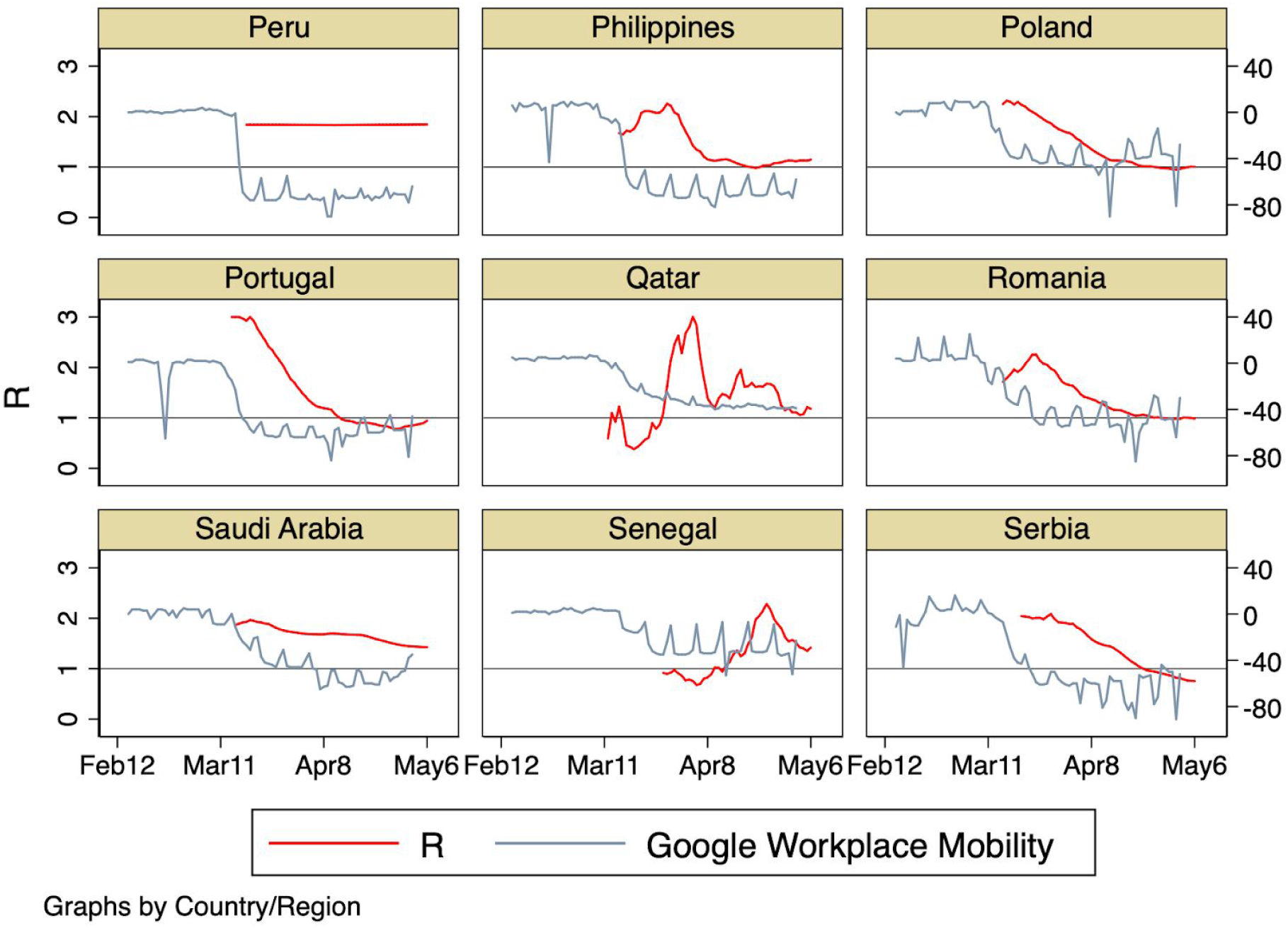
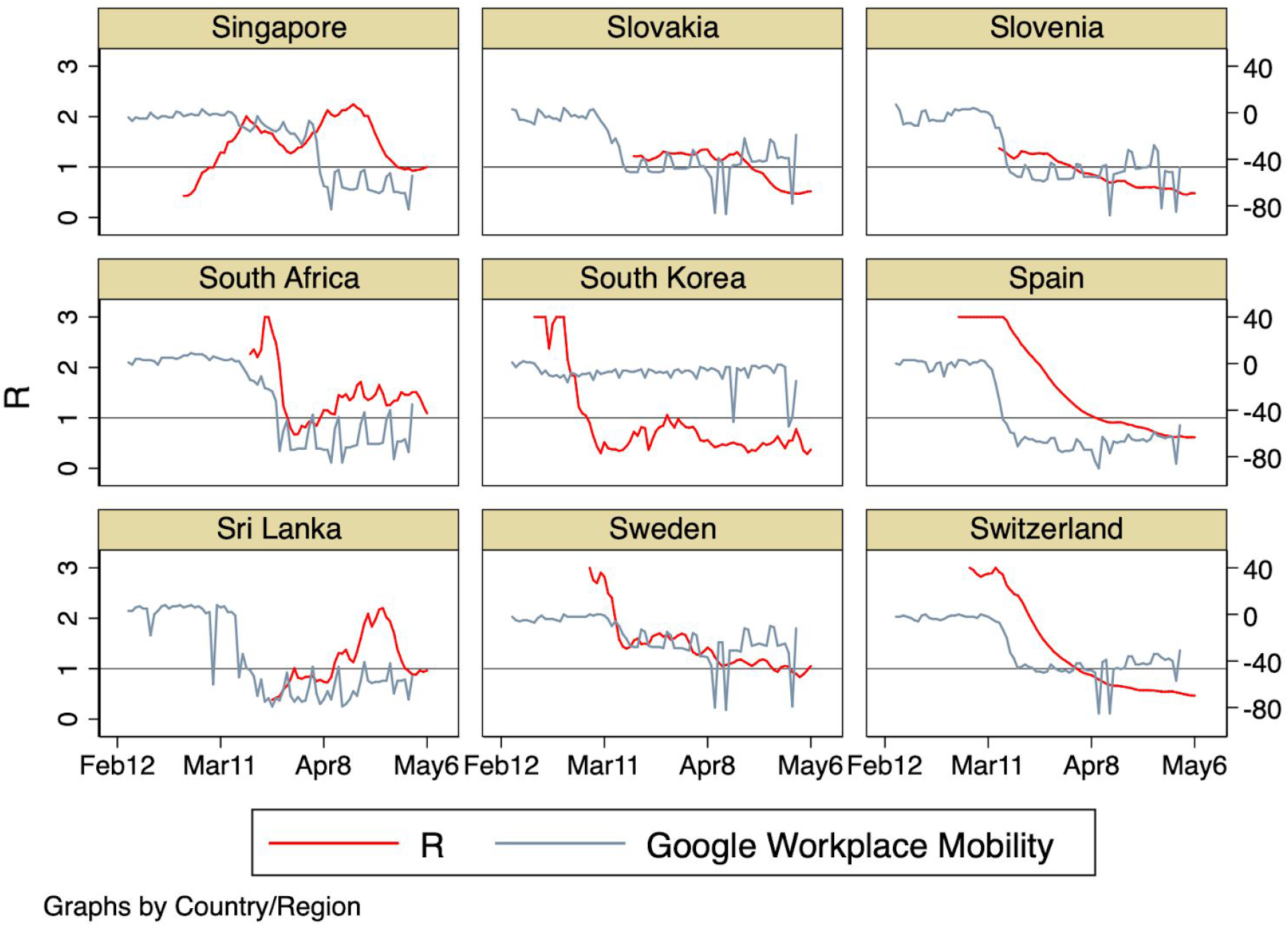
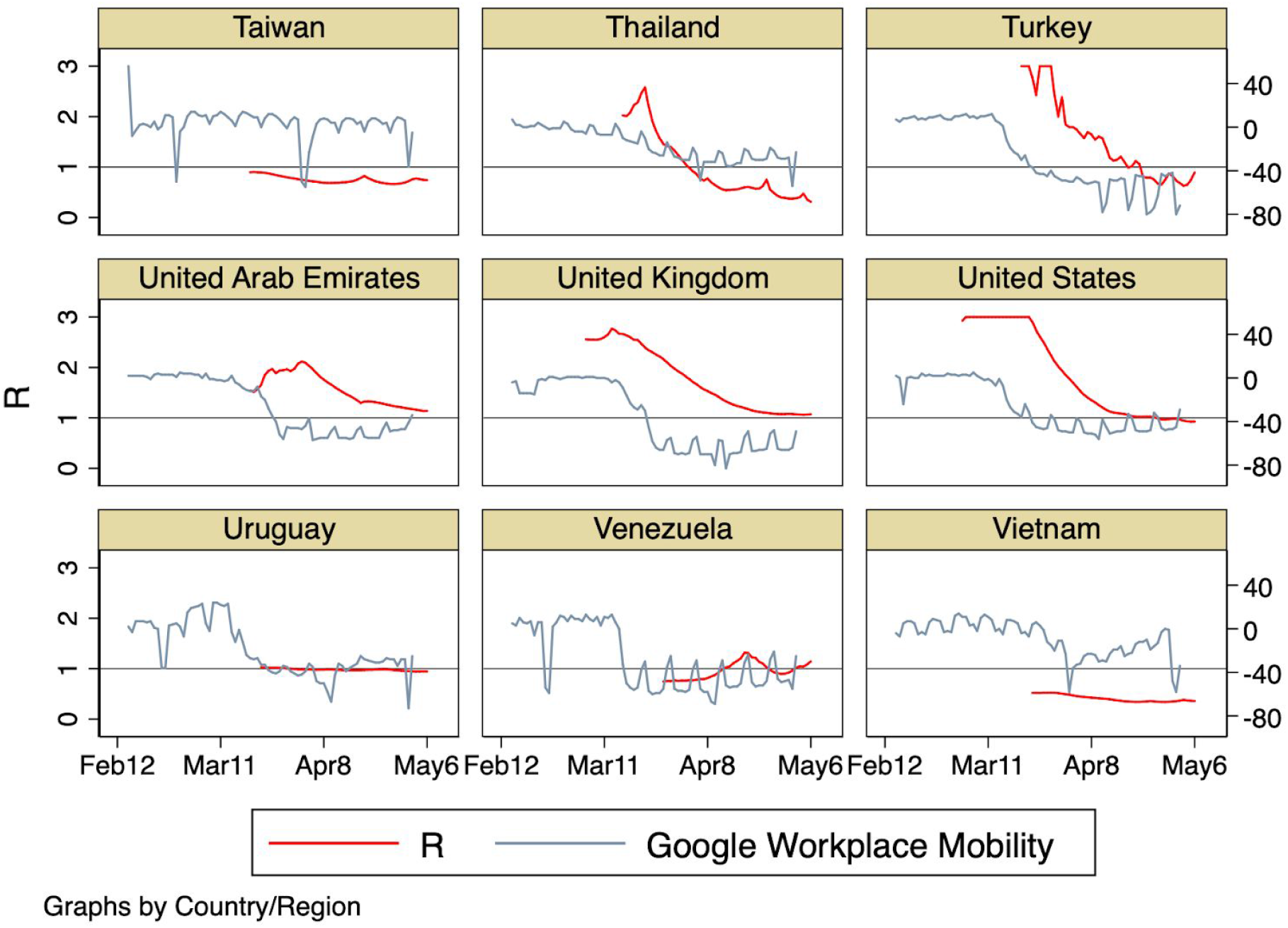
Country plots of *R*(*t*) and Google workplace mobility over time. Only countries for which there are at least 30 days of both mobility and *R*(*t*) data are included. *R*(*t*) is plotted against the left axis. Mobility changes from baseline (in percentage points, see datasection) are plotted against the right axis.

**Figure S5:**
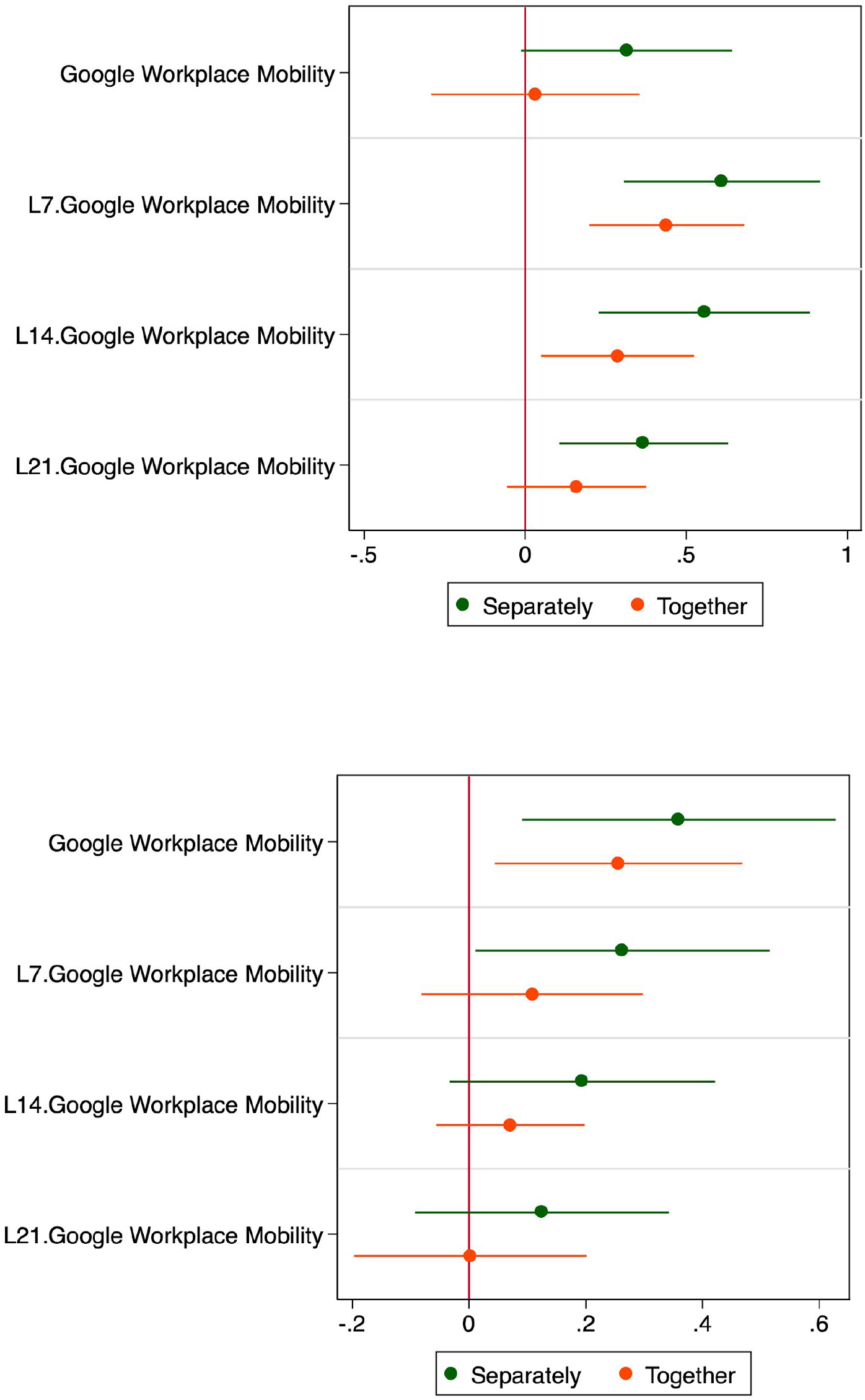
Coefficients of regressions of estimated *R*(*t*) values on various lags of the Google workplace mobility indicator as the explanatory variable, estimated in separate regressions (green markers) or jointly (orange markers) for the international panel (top) and the U.S. state panel (bottom). Error bars indicate 95% confidence intervals. All regressions include country/state and date fixed effects.

**Figure S6:**
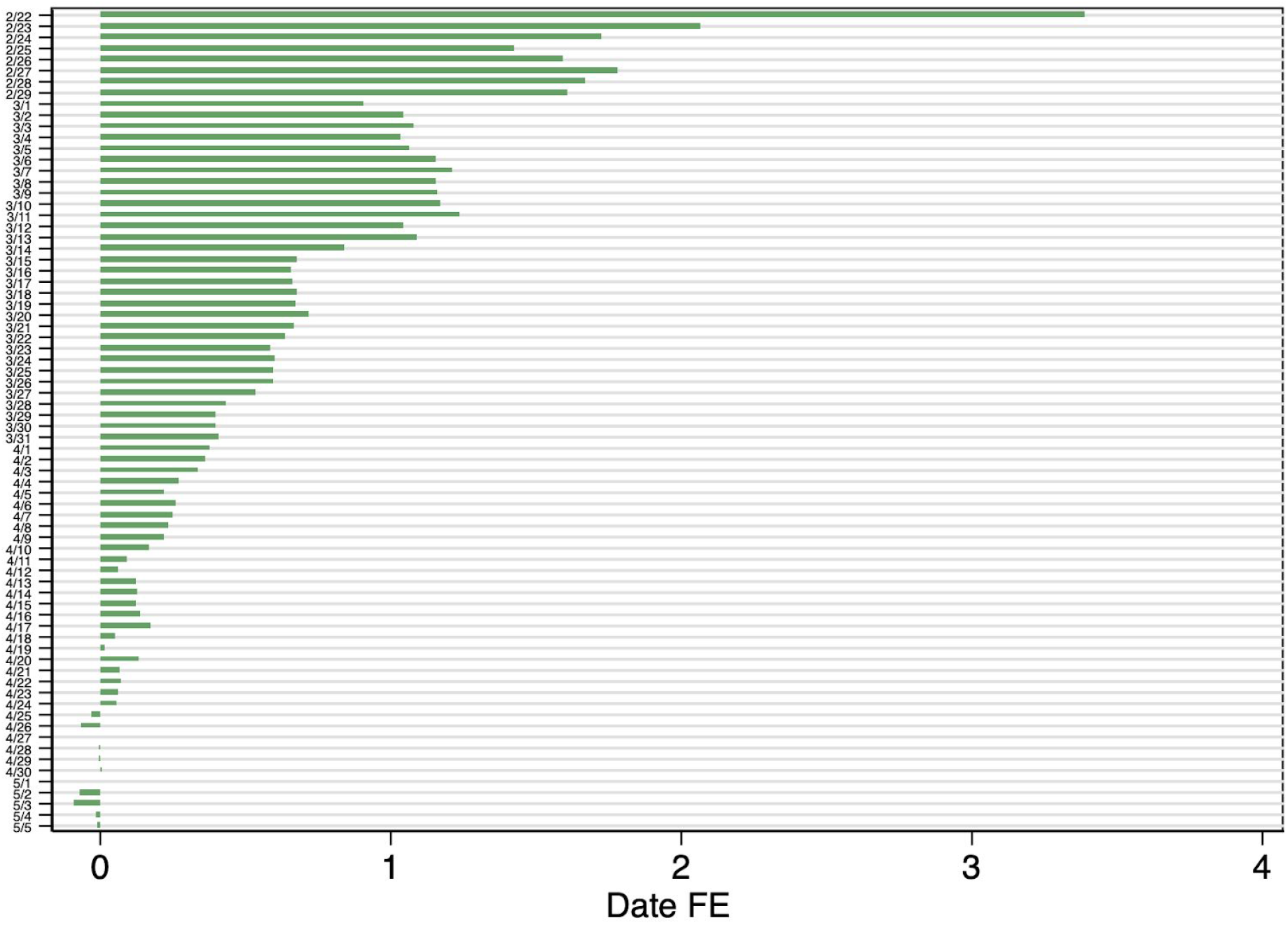
Estimated date fixed effects from regression (1), sorted chronologically.

**Table S1:**
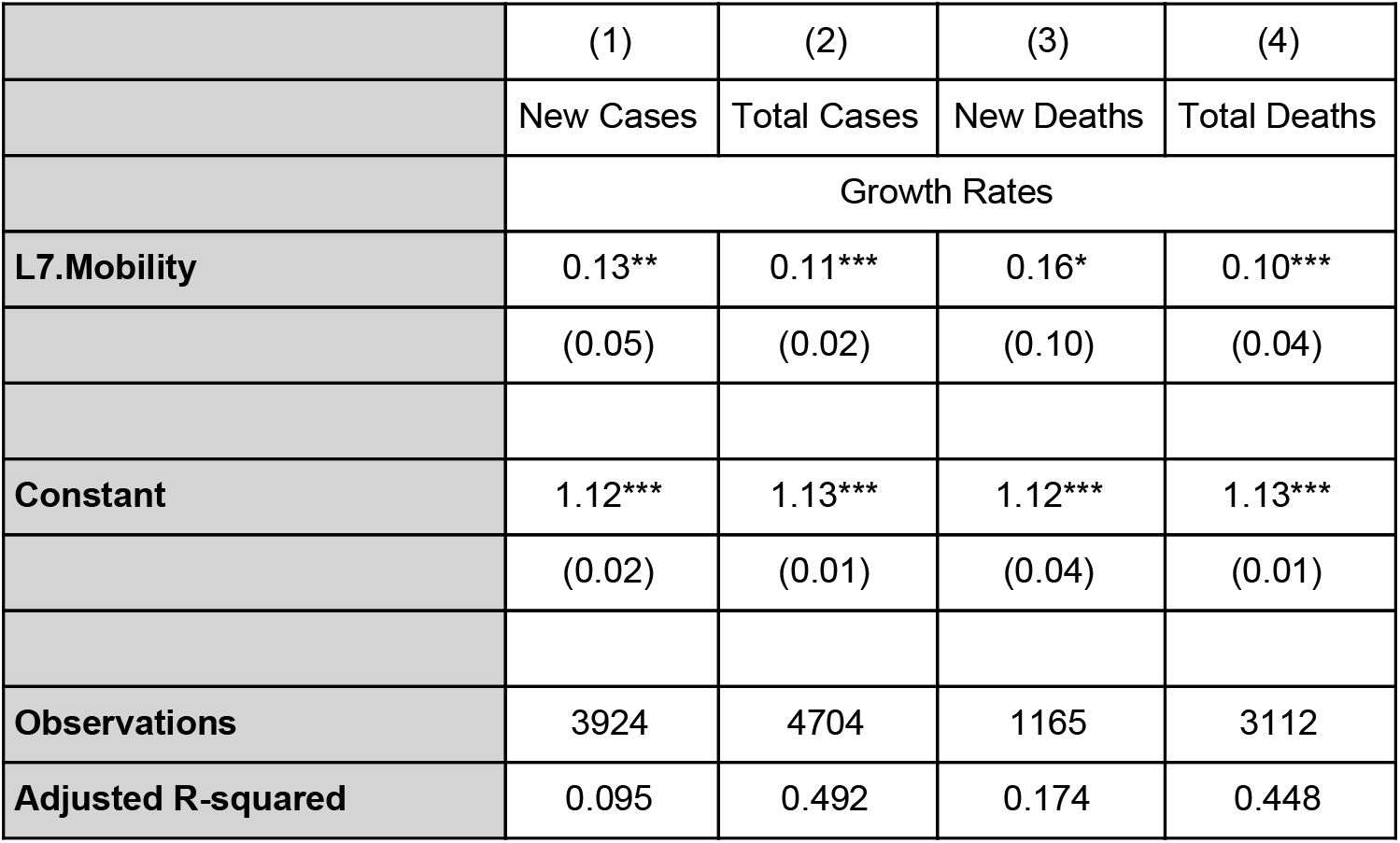
Regression results relating alternative measures of Covid-19 transmission to the 7-day lagged Google workplace mobility indicator. Column 1: dependent variable is the daily growth rate of new cases (3-day running mean). Column 2: dependent variable is the daily growth rate of total confirmed cases. Column 3: dependent variable is the daily growth rate of new deaths (3-day running mean). Column 4: dependent variable is the daily growth rate of total deaths. All regressions include country and date fixed effects. Standard errors, clustered by country, are reported in parentheses. Stars indicate statistical significance (*p<0.1, **p<0.05, ***p<0.01).

## Footnotes

1 We thank Yinon Bar-On, Joshua Graff Zivin, Joseph Klafter, Ofer Malamud, Ron Milo, Zvi Mintser, Jeffrey Sachs, Itay Saporta, Analia Schlosser, and Anna Tompsett for useful comments and suggestions.

4 Available at rt.live.

5 As one example, government ordered lockdowns may have acted as a signalling device indicating the severity of the health crisis, thereby influencing individual behavior.

6 These include non-mobility related social distancing and other non-pharmaceutical interventions (NPIs), or potentially, through environmental changes such as warming or increased UV radiation (Carleton et al, 2020) which may independently also reduce transmission rates.

7 These estimates are calculated from the analysis conducted on the full sample period. To the extent that there are interaction effects between various methods of transmission suppression — for example, if mask usage reduces the impact of increased mobility on *R* — required reductions in transmission rates will be lower. This issue is the subject of ongoing work.

8 The baseline period is the median value for the corresponding day of week, calculated during the 5-week period Jan 3–Feb 6, 2020.

9 https://www.google.com/Covid19/mobility/

10 Apple data extended beyond Google data at the day of data download.

11 Qun Li et al. 2020 report an average incubation period of 5 days and Linton et al. report a median incubation period of 4–5 days.

12 For the state-level analysis in the United States, all regressions include state-level fixed effects.

13 We classify countries into six geographical regions: Africa, Asia and Pacific, Europe, Middle East, North America and South/Latin America.

14 For example, a 10 percentage point reduction in mobility is coded as -0.1.

15 The omitted regional fixed effect is Europe.

16 As there are only two countries in the North America region in our data, we do not run the regression separately for that region.

17 To estimate R, Kučinskas (2020) requires that the number of reported Covid-19 cases exceed 100, implying that countries where infection rates climbed in later time periods are underrepresented in the sample.

18 Given the assumption of a seven-day lag between actual and observed R in the international data, the values corresponding to a given date refer to the R estimated seven days afterwards.

